# A cystic fibrosis lung disease modifier locus harbors tandem repeats associated with gene expression

**DOI:** 10.1101/2022.03.28.22272580

**Authors:** Delnaz Roshandel, Scott Mastromatteo, Cheng Wang, Jiafen Gong, Bhooma Thiruvahindrapuram, Wilson W.L. Sung, Zhuozhi Wang, Omar Hamdan, Joe Whitney, Naim Panjwani, Fan Lin, Katherine Keenan, Angela Chen, Mohsen Esmaeili, Anat Halevy, Julie Avolio, Felix Ratjen, Juan C. Celedón, Erick Forno, Wei Chen, Soyeon Kim, Lei Sun, Johanna M. Rommens, Lisa J. Strug

## Abstract

Variable number of tandem repeats (VNTRs) are major source of genetic variation in human. However due to their repetitive nature and large size, it is challenging to genotype them by short-read sequencing. Therefore, there is limited understanding of how they contribute to complex traits such as cystic fibrosis (CF) lung function. Genome-wide association study (GWAS) of CF lung disease identified two independent signals near *SLC9A3* displaying a high density of VNTRs and CpG islands. Here, we used long-read (PacBio) phased sequence (N=58) to identify the boundaries and lengths of 49 common (frequency >2%) VNTRs in the region. Subsequently, associations of the VNTRs with gene expression were investigated in CF nasal epithelia using RNA sequencing (N=46). Two VNTRs tagged by the two GWAS signals and overlapping CpG islands were independently associated with *SLC9A3* expression in CF nasal epithelia. The two VNTRs together explained 24% of *SLC9A3* gene expression variation. One of them was also associated with *TPPP* expression. We then showed that the VNTR lengths can be estimated with good accuracy in short-read sequence in a subset of individuals with data on both long (PacBio) and short-read (10X Genomics) technologies (N=52). VNTR lengths were then estimated in the Genotype-Tissue Expression project (GTEx) and their association with gene expression was investigated. Both VNTRs were associated with *SLC9A3* expression in multiple non-CF GTEx tissues including lung. The results confirm that VNTRs can explain substantial variation in gene expression and be responsible for GWAS signals, and highlight the critical role of long-read sequencing.

## INTRODUCTION

Variable number of tandem repeats (VNTRs) with repeat units ≥ 6 base pairs (bps) often span hundreds of bps and are major sources of variation in humans. However, due to their repetitive nature and large size, it is challenging to genotype them by short-read sequencing. This technological obstacle has limited our understanding of how they contribute to complex traits ^1^. Targeted studies have shown that variation in VNTR length can affect disease phenotypes. For instance, a 14-mer VNTR upstream of *INS* (insulin) has been associated with multiple metabolic traits and diseases including diabetes, obesity and polycystic ovary ^2^; a 25-mer intronic VNTR within *ABCA7* (ATP binding cassette subfamily A member 7) has been associated with Alzheimer disease ^3^; and a VNTR in the first exon of *AS3MT* (arsenite methyltransferase) has been associated with schizophrenia ^4^. Additionally, variation in the length of many VNTRs located in coding regions, untranslated regions (UTRs) and regulatory regions proximal to genes has been associated with nearby gene expression (eVNTRs) ^1; 5^ and DNA methylation levels (mVNTRs) ^5^. We were interested in the extent to which VNTRs explain the contribution of a complex locus the subtelomeric proximity of chromosome 5p to Cystic Fibrosis (CF) lung disease, identified in the largest genome-wide association study (GWAS) of CF by the international CF gene modifier consortium ^6^.

CF is a life-limiting autosomal recessive disease affecting ∼70,000-100,000 individuals globally^7^. It is caused by variations in the cystic fibrosis transmembrane conductance regulator (*CFTR*; OMIM:602421) with p.Phe508del (c.1521_1523delCTT) being the most frequent ^8; 9^. However, to date, over 2,100 different variations have been reported in the gene (Cystic fibrosis mutation database, accessed March 2, 2021), with 431 of them demonstrated to cause CF (CFTR2 website, accessed November 4, 2021) ^10^. CFTR is located in the apical membrane of epithelial cells in many tissues ^11^.

CF affects multiple organs including the lungs, sweat glands, liver, pancreas, intestine and vas deferens of the male reproductive tract. However, progressive chronic obstructive lung disease is responsible for the majority of morbidity and mortality ^11^, and there is a high degree of variability in disease severity, complications and survival among individuals with CF. This variability can be explained by different *CFTR* mutations ^12–14^, the environment ^15^ and gene modifiers ^6; 16–19^. Family and twin studies have shown that about half of the variation in lung function, measured with spirometry or by susceptibility to infection with *Pseudomonas aeruginosa* (PsA), can be attributed to genetic modifiers ^20–22^. Several gene modifier loci have now been shown to contribute to disease variability in CF lungs ^6; 23; 24^.

A meta-GWAS of lung function in 6,365 individuals with CF identified five genome-wide significant loci including the sub-telomeric locus on chromosome 5 that contains solute carrier family 9 member A3 (*SLC9A3*) ^6^, the same locus that had previously been identified in a GWAS of CF intestinal obstruction ^16^. Lung function was defined as average (over 3 years) age-specific CF percentile values of forced expiratory volume in one second (FEV1) for each patient relative to other CF patients of the same age, sex and height, and adjusted for mortality ^6^. The lung function locus contained two independent association signals confirmed by conditional analysis, rs57221529 (CHR5:586,509; A>G) 5’ of *SLC9A3* and rs72711364 (CHR5:415,208; A>G) 3’ of *SLC9A3* [Figure S1] ^6^. A SNP in this region, rs11738281 (CHR5: 662,432; C>T), in high linkage disequilibrium (LD) with rs57221529 (R^2^ = 0.76, D’ = 0.90) has also been reported to associate with lung function in non-CF individuals (age range between 40 and 69 years) from the UK Biobank ^25^.

SLC9A3 is an epithelial Na^+^/H^+^ exchanger expelling acid from the cell using an inward sodium ion gradient. Although *SLC9A3* is expressed in lung epithelia according to Human Protein Atlas, functional investigations have focused on other organs. *SLC9A3* deficiency causes congenital secretory sodium diarrhea ^26^. *Slc9a3*^-/-^ mice experience diarrhea and acidosis due to reduced fluid absorption in the intestine and proximal convoluted tubules, respectively ^27^. Males also develop obstructive azoospermia due to abnormal abundant secretions and calcification in their reproductive tract lumen ^28^. *Cftr*-null mice lacking one or both copies of *Slc9a3* also demonstrate increased fluidity in their intestinal contents with at least partial resolution of the obstructions commonly seen in CF mouse models ^29^.

While *SLC9A3* provides an interesting candidate gene, GWAS identify loci rather than causal genes. In addition to *SLC9A3*, aryl-hydrocarbon receptor repressor (*AHRR*), exocyst complex component 3 (*EXOC3*), centrosomal protein 72 (*CEP72*), and tubulin polymerization promoting protein (*TPPP*) are also annotated to the locus (CHR5:403,462-686,129 (HG38)). With the exception of *AHHR*, all of these genes are expressed in lung epithelia according to Human protein atlas. EXOC3 is a component of the exocyst complex, a multiple protein complex essential for targeting exocytic vesicles to specific docking sites on the plasma membrane. Transport vesicle processes have been significant in CF gene expression studies and GWAS pathway analysis ^30; 31^. CEP72 and TPPP both function in microtubule formation. Microtubules are part of the cytoskeleton that provide a platform for intracellular transport including movement of secretory vesicles, and their formation is disrupted in CF cells ^32^. Thus, a thorough investigation of the locus to determine the responsible gene and causal variation marked by the GWAS signal is required.

None of the genome-wide significant SNPs in the region are, or are in LD with, protein coding variants, suggesting that the causal variation acts through other mechanisms such as gene expression alteration. Based on the Genotype-Tissue Expression (GTEx) project, rs57221529 is an expression quantitative trait locus (eQTL) for all five genes; and rs72711364 is an eQTL for all but *TPPP* in multiple tissues [Table S1 & S2]. The associated locus displays an exceptionally high density of CpG islands (GC content ≥50%, length >200 bp, and ratio >0.6 of observed number of CG dinucleotides to the expected number on the basis of G and C nucleotide content ^33^). Chromosome 5 on average includes nine CpG islands per 1 Mb (1 per gene) ^34^, while this 280 kb locus includes 33 CpG islands based on UCSC. Furthermore, methylation levels at CpGs in this region have been associated with *SLC9A3* expression in nasal epithelia of individuals without CF ^35^. Another notable feature of this locus is that it includes a large number of VNTRs^36^. The exact lengths and repeat units of these VNTRs have not been well defined. Both features -- high numbers of CpGs and VNTRs -- are expected as the locus partly overlaps the subtelomeric region (500 kb of each autosomal arm terminal) which is known to have a high density of DNA repeats and CpG sequences ^37^.

Here, we hypothesize that the VNTRs tagged by GWAS SNPs in the region affect lung function in individuals with CF by altering expression of nearby genes through changing methylation levels at CpGs.

First, we test this indirectly. We use DNA methylation quantitative trait loci (mQTL) and expression quantitative trait DNA methylation (eQTM) data from non-CF nasal epithelia. We show that the same GWAS SNPs that are associated with CF lung function are also associated with methylation levels at nearby CpGs, whose methylation levels associate with *SLC9A3* expression. Integrating long-read sequencing and phasing along with RNA-sequencing of nasal epithelia from individuals with CF, we identify eVNTRs in the region that are tagged by the GWAS and mQTL SNPs. Finally, we estimate the eVNTR lengths in GTEx short-read sequence to extend the VNTR association with gene expression in non-CF lung tissue as well as multiple other GTEx tissues [Figure 1].

**Figure 1:**
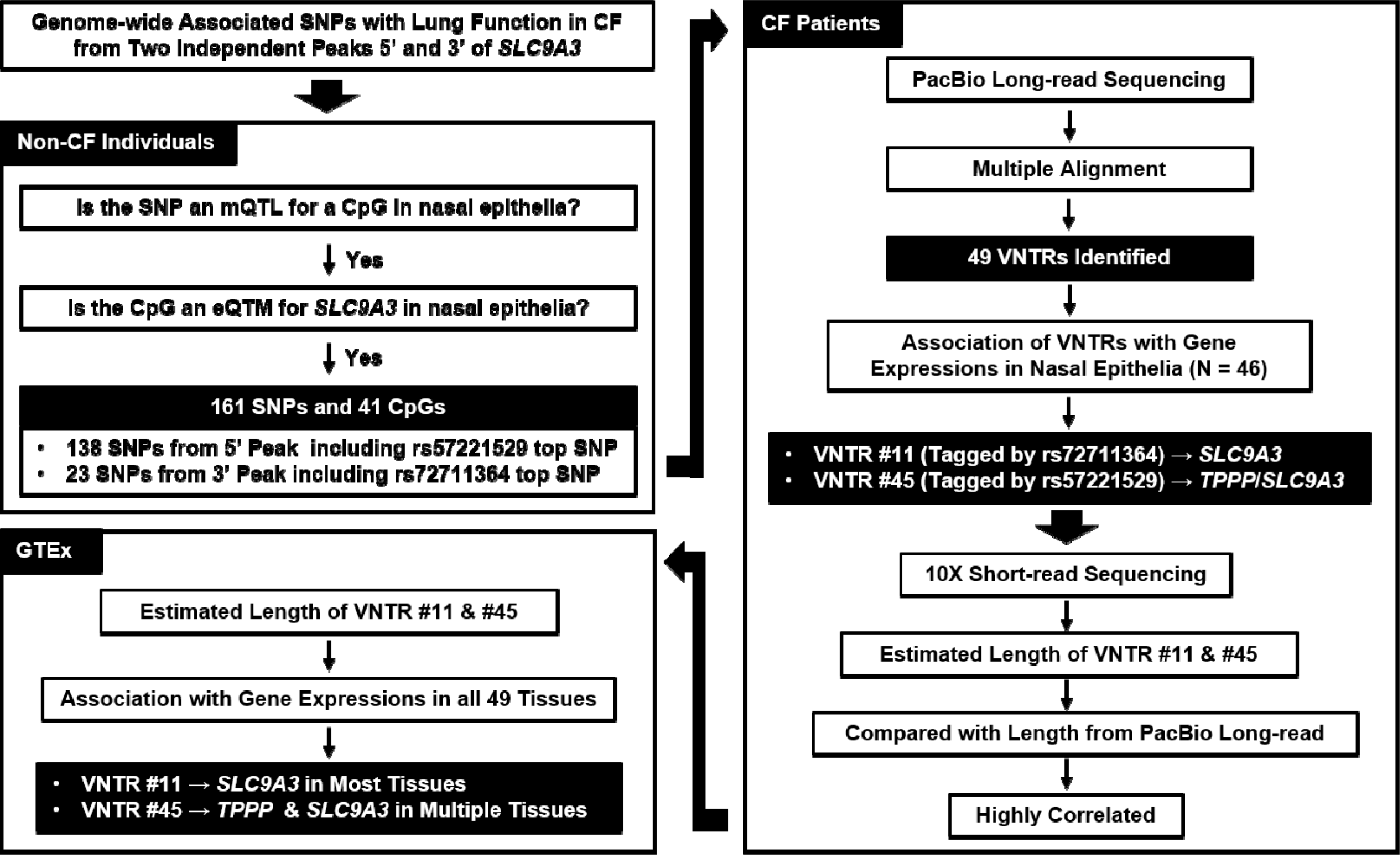
Study design and main findings

## MATERIAL AND METHODS

### Non-CF nasal epithelia mQTL and eQTM analysis

Whole-genome mQTL and eQTM data from the nasal tissue in 455 non-CF subjects (with or without asthma) ^35^ were used to investigate if any of the previously identified SNPs associated with lung function at the genome-wide significance threshold ^6^ are mQTLs for nearby (±100Kb) CpGs in *SLC9A3* ± 1 Mb region (false discovery rate (FDR) p <0.05) that affect *SLC9A3* expression in the nasal tissue (p <0.05). Whole-genome DNA methylation assessment was done using HumanMethylation450 BeadChips (Illumina, San Diego, CA, USA). All mQTL models were adjusted for asthma and atopy status, age, sex, methylation batches, and a latent factor of methylation calculated from the sva R package. RNA-Seq was done with the Illumina NextSeq 500 platform (Illumina; San Diego, CA, USA). Asthma and atopy status, age, sex, the top five principal components (PCs) from genotypic data, RNA cell type (sorted or not), methylation and RNA-Seq batch, and latent factors of methylation and RNA-seq (estimated using the R package sva) were included as covariates in the eQTM analysis ^35^.

### CF Subjects

Subjects with CF were recruited into the Canadian Cystic Fibrosis Gene Modifier Study (CGMS) from 35 sites across 9 Canadian provinces. CGMS was approved by the Research Ethics Board (REB) of the Hospital for Sick Children (# 0020020214 from 2012-2019 and #1000065760 from 2019-present), and by the respective REBs at each of the other participating sites. Written informed consent was obtained from all participants or their parents/guardians/substitute decision makers. Long-read sequencing Pacific Biosciences (PacBio) was performed for a subset (N = 58) of CGMS participants. A larger subset (N = 447) of CGMS subjects had their DNA sequenced using the linked-read technology from 10X Genomics (10XG), a subset (N = 52) of which overlap with those sequenced using PacBio. Nasal epithelia tissue was also collected from a subset (N = 131) of CGMS samples and bulk RNA sequenced. Of these 131 subjects, 46 also had PacBio and 68 also had 10XG [Figure 1]. Fifty-one percent of the participants included in the current study were homozygous for p.Phe508del, 18% were heterozygous for p.Phe508del, and 31% had two other causal variants.

### DNA extraction

High molecular weight (HMW) DNA was extracted from fresh or frozen blood aliquots using MagAttract HMW DNA kit (QIAGENE, Cat# 67563) as per supplier recommendations. Quantitation was determined by Quant-iT PicoGreen dsDNA assay Kit (Invitrogen, Cat# P11469) as recommended by the supplier. Quality of DNA was also assessed by electrophoretic migration in 0.4% agarose gel, run at 50 Volt for 18 hours at 4 ◦C in Tris-acetate buffer at pH 8.0 with comparison to Quick load 1kb extend DNA ladder (NEB, Cat# N3239S). Samples with bulk DNA > 50kb (>80% by visual inspection of agarose gel) were submitted for sequencing.

### PacBio long-read sequencing

About 10-15 μg of genomic DNA was submitted to The Centre for Applied Genomics (TCAG), the Hospital for Sick Children, Toronto, Canada for genomic library preparation and whole genome sequencing. DNA samples were quantified using Qubit dsDNA High Sensitivity Assay and sample purity was checked using NanoDrop OD 260/280 ratio. DNA samples were run on the Genomic Tape on TapeStation (Agilent, Cat# 5067-5365 and 5067-5366) to check if DNA is of high molecular weight (>60 Kb, DNA integrity number (DIN) >7). DNA was sheared to about 20-30 Kb using Covaris g-TUBEs, quantified using Qubit High Sensitivity Assay and checked on the Genomic Tape on Tapestation for sheared fragment size. About 5 μg of DNA was used as input material for library preparation using the SMRTbell Express Template Prep Kit 2.0 following the manufacturer’s recommended protocol. In brief, DNA was end-repaired, ligated with blunt-end adapters, and size-selected using the BluePippin System to remove fragments below the desired size of 20kb; libraries were validated on the Genomic Tape on Tapestation to check for size and quantified by the Qubit High Sensitivity Assay. Validated libraries were set up for sequencing on the PacBio Sequel II following PacBio’s recommended protocol; libraries were primer-annealed and polymerase-bound for Diffusion loading on two 8M SMRT Cells per sample for a maximum movie time of 15 hours per SMRT Cell.

PacBio reads were aligned using the PacBio wrapper for minimap2, pbmm2 v1.3.0.

### 10XG linked-read sequencing

Approximately 1 μg of genomic DNA was submitted to TCAG for genomic library preparation and whole genome sequencing. DNA samples were quantified using Qubit dsDNA High Sensitivity Assay and sample purity was checked using NanoDrop OD 260/280 ratio. DNA was run on the Genomic Tape on TapeStation (Agilent, Cat# 5067-5365 and 5067-5366) to check DNA fragment size. DNA with DIN >7 proceeded to the next step. DNA was damage repaired using the PacBio SMRTbell Damage Repair Kit-SPv3 (PN 100-992-200). Subsequently, it was cleaned using Pacbio Ampure PB magnetic beads (PN 100-265-900). About 10 ng of damage-repaired DNA was used as input material for library preparation using the 10x Library Kit (PN 120258 and 120257) following the manufacturer’s recommended protocol. Libraries were validated on a Bioanalyzer DNA High Sensitivity chip to check for size and absence of primer dimers; and quantified by qPCR using Kapa Library Quantification Illumina/ABI Prism Kit protocol (KAPA Biosystems). Validated libraries were sequenced on an Illumina HiSeq X platform following Illumina’s recommended protocol to generate paired-end reads of 150-bases in length.

Long Ranger v2.2.2 ^38^ was used to align 10x reads to HG38.

### RNA-seq

Extraction was performed via nasal brushing on either inferior turbinate using a 3-mm diameter sterile cytology brush (MP Corporation, Camarillo, CA) or Rhino-probe curette. RNA was extracted using Qiagen RNeasy Micro kit. Bulk RNA sequencing was performed on the HNE samples in two batches. The first batch was sequenced on the Illumina HiSeq 2000 platform (Illumina Inc. San Diego, California, USA) with the specification of 25 million paired-end 49 base pair in length reads per sample. The second batch was sequenced using the Illumina HiSeq 2500 platform with high output mode flow cell and V4 chemistry. Each sample in the second batch was sequenced with 35 million paired-end 124 base pair reads.

Trimming of the raw sequenced reads was carried out using Trim Galore v0.4.4. STAR (ver. 2.5.4b) ^39^ was used to align the processed reads to human reference genome HG38 in conjunction with GENCODE comprehensive gene annotation (release 29). Read counts per gene were quantified using RNA-SeQC (ver. 2.0.0) and normalized to transcripts per million (TPM)^40^. Normalized trimmed mean of M values (TMM) measures were generated using genes with ≥0.1 TPM and ≥6 read counts in ≥20% of the samples^41^.

### Identifying VNTR boundaries and lengths

We identified VNTRs using reference-aligned PacBio reads. For each of our PacBio samples, we isolated the region of interest where CF lung function genome-wide significant SNPs are located (p <5×10^-8^) extended on both ends with an additional 10kb of flanking sequence (Chr5:393,462-6896,129, HG38). GCpp v1.9.0 was used to polish the reference sequence using the aligned PacBio reads (using a large chunk size -C 5000). PacBio reads were realigned to the polished consensus sequence using pbmm2 v1.3.0. The new alignments and Longshot v0.4.1 ^42^ were used to identify heterozygous SNPs and to phase the PacBio reads into two haplotypes. Finally, an iterative polishing approach was applied separately to each haplotype: the reference sequence was polished using GCpp, and the haplotype-specific reads were realigned to the polished output. This iterative approach generates a series of FASTA files that converges to a consensus of the haplotype-specific reads. In short, a consensus sequence for each haplotype is generated. We found that two iterations of polishing and alignment was sufficient for convergence. Subsequently, we used MAFFT ^43^ to perform multiple sequence alignment of all haplotypes. VNTR (repeats with repeat units ≥6 bp) boundaries were defined by polymorphic regions with repeat structures. Subsequently, the exact length of each VNTR was calculated for both haplotypes of all 58 subjects with CF.

Spearman correlation was used to investigate the correlation between lengths of different VNTRs, and between number of CpGs in VNTRs and their corresponding lengths.

### VNTR associations with gene expressions in CF nasal epithelia

Linear regression was used to test the association of VNTR lengths/number of CpGs (mean of the two haplotypes) and gene expression (TMM) in 46 CF patients with both PacBio and RNA-seq. Sex, RNA integrity number (RIN), first three PCs, seven first probabilistic estimation of expression residuals (PEER) factors and *CD45* expression (as a proxy for immune cells aggregation in nasal tissue due to infection) ^44^ were included in the regression model as covariates.

Matrix Spectral Decomposition (matSpD) method was used to calculate the number of independent VNTRs in the region ^45^.

Since most of pseudo-genes and non-coding RNAs did not meet the inclusion criteria (i.e. ≥0.1 TPM and ≥6 read counts in ≥20% of the samples), they were not included in the current analysis.

### SNP associations with gene expression in CF nasal epithelia

The associations of rs57221529 and rs72711364, the top SNPs from the GWAS peak 5’ and 3’ of *SLC9A3*, with gene expression were tested in 68 subjects with both 10XG and RNA-seq data using linear regression with sex, RIN, first three PCs, seven first PEER factors, and *CD45* expression included in the model as covariates.

### Estimating VNTR lengths in short-read sequence data

The lengths of associated VNTRs with gene expressions in nasal epithelia (mean of the two haplotypes) were estimated in 10XG short-read sequencing by dividing the number of all reads aligned to the location of each VNTR by the average sequencing depth in 52 CF subjects with both PacBio and 10XG sequencing. Spearman correlation was used to investigate the concordance between VNTR lengths in PacBio and their estimated lengths in 10XG in participants with data from both technologies.

### GTEx V8

BAM files from whole genome sequencing in 646 subjects were downloaded from dbGaP accession number phs000424.v8.p2 on Sep 2020. Subsequently, all reads aligning to the location of associated VNTRs with gene expression in CF nasal epithelia were extracted. Then, VNTR lengths were estimated by dividing the number of reads aligned to the location of each VNTR by the average sequencing depth.

Fully processed, filtered and normalized gene expression matrices (in BED format) for each tissue as well as covariates used in eQTL analysis were downloaded from the GTEx portal. Linear regression was used to test the association between the estimated VNTR lengths and gene expression in all 49 tissues. Top five genotyping PCs, PEER factors determined by sample size (15 factors for N <150, 30 factors for 150≤N<250, 45 factors for 250≤N<350, and 60 factors for N≥350), sequencing platform, sequencing protocol, and sex were included in the regression model as covariates.

### *SLC9A3* expression in CF vs. non-CF nasal epithelia

Linear regression was used to compare *SLC9A3* expression in CF vs. non-CF nasal epithelia by active PsA infection status (confirmed by sputum culture within 2 weeks from RNA sampling). RIN, study site, first three PEER factors, and *CD45* expression were included as covariates in the regression model.

## RESULTS

### Integrating CF lung function GWAS with non-CF methylation and expression in nasal epithelia

To determine whether the GWAS association signal at the *SLC9A3* locus could be due to changes in gene expression as a result of DNA methylation, we initially integrated mQTL ^46^ and eQTM ^47^ summary statistics in non-CF nasal epithelia ^35^ with the CF lung function GWAS summary statistics ^6^. Of the 212 genome-wide significant CF lung function associated SNPs (CHR5: 403,462-686,129), 161 were associated with methylation levels at 41 CpGs (CHR5:467,037-664,299) that in turn were associated with *SLC9A3* expression [Figure 2, Supplementary Excel file]. The pairwise LD between these SNPs demonstrates that they belong to two LD blocks tagged by the two independent GWAS peaks, one positioned 5’ and the other 3’ of *SLC9A3* (oriented on the reverse strand). The 5’ block comprises 138 SNPs (CHR5:510,378-686,129) from *TPPP* to intron 1 of *SLC9A3* including rs57221529, the top CF lung function GWAS SNP annotated to this 5’ peak. These 138 SNPs were associated with methylation levels at CpGs spanning *TPPP* to *SLC9A3*. The 3’ block includes 23 SNPs (CHR5:403,462-476,238) from exon 13 of *SLC9A3* to *AHRR* including rs72711364, the top CF lung function GWAS SNP from this peak. These 23 SNPs were associated with methylation levels at CpGs positioned from *SLC9A3* to *EXOC3*. There were ten CpGs spanning *SLC9A3* where SNPs from both the 5’ and 3’ LD block were associated with their methylation levels [Figure S2]. These findings suggest that the GWAS association could be due to *SLC9A3* gene expression variation, but the complex sequence architecture at the locus which includes many VNTRs makes it difficult to determine the causal genetic variation.

**Figure 2:**
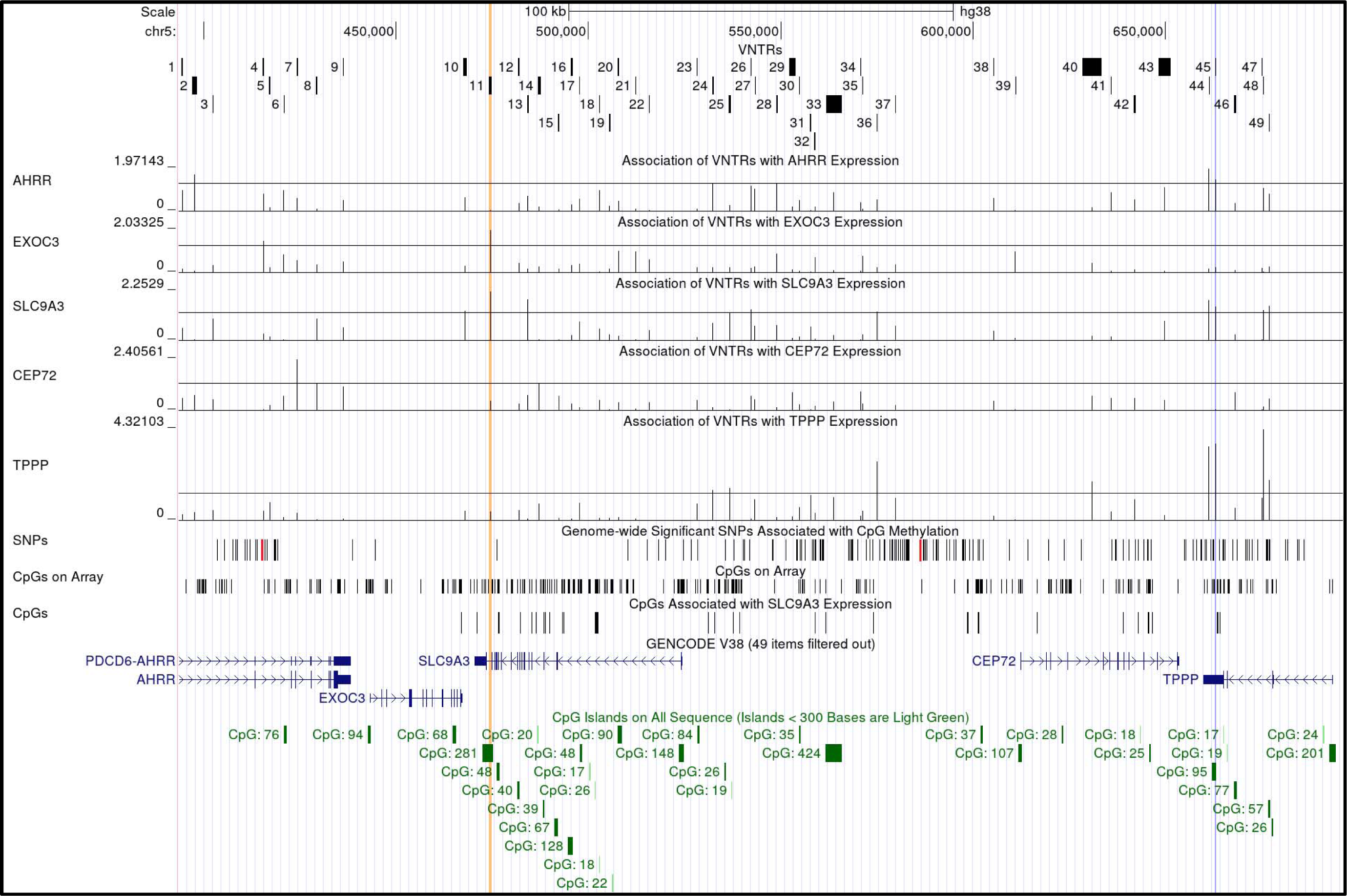
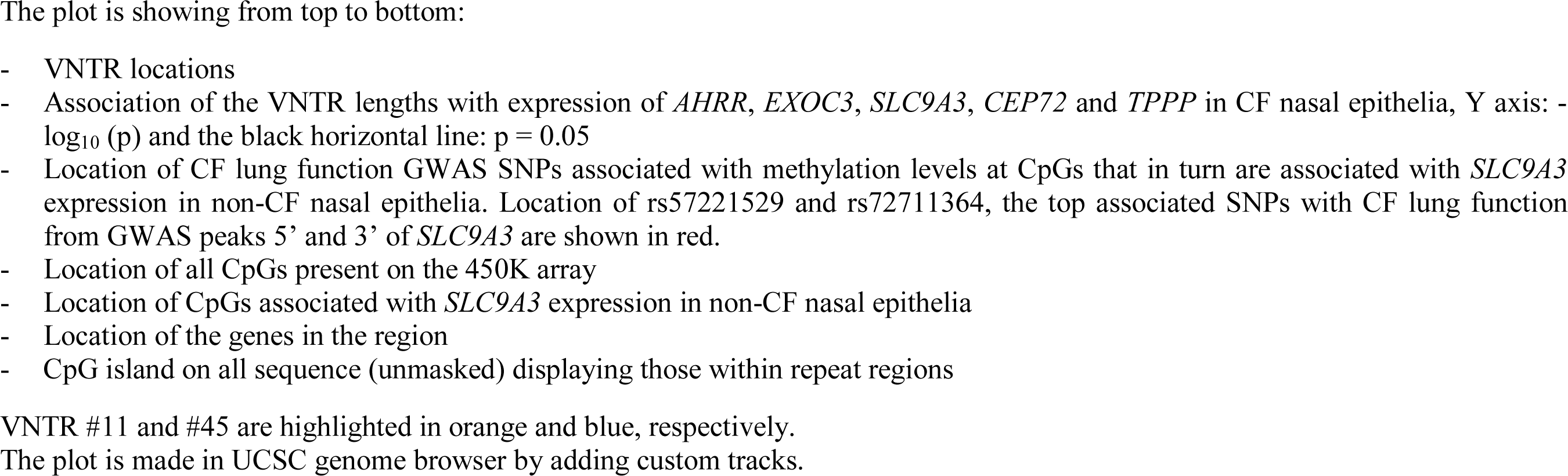
Association of VNTRs with gene expressions in CF nasal epithelia, and location of CF lung function GWAS SNPs associated with methylation levels at CpGs that in turn were associated with *SLC9A3* expression in non-CF nasal epithelia

### Identifying VNTR boundaries and their lengths using PacBio long-read sequence

Long-read phased data from PacBio whole genome sequencing technology was used to identify the VNTR boundaries and lengths. The consensus sequences of the two haplotypes for all individuals plus the reference human genome (HG38) were subjected to multiple alignment to identify the regions with variable lengths and repeat structure corresponding to the polymorphic parts of the VNTRs (as outlined in the Methods section). We identified a total of 49 VNTRs across the region plus its 10kb flanking sequence (Chr5:403,462-686,129 ± 10kb). Large segmental duplications (>45 kb) that could not be phased are annotated to the end of the region (Chr5:677,960-696,129), but the CF lung function association pattern does not implicate this region as contributing ^6^. All but VNTR #1, 39 and 47, contain CpGs; and 14 of the VNTRs overlap with CpG islands based on UCSC genome browser annotation of CpG islands ^33^ [Table 1, Figure 2]. For VNTRs containing CpGs, the number of CpGs was positively correlated with the VNTR length [Table S3] and the length of some VNTRs were correlated with each other [Table S4]. We calculated 46 independent VNTRs in the region and used a regional significance threshold of 0.001 (0.05/46).

**Table 1:** Characteristics of the common VNTRs identified in the region (CHR5: 403,462-686,129 ± 10kb)

| VNTR | Start | End | CpG<br>Island<br>Overlap | HG38 | Length (bp) |  |  | HG38 | CpGs (N) |  |  |
| --- | --- | --- | --- | --- | --- | --- | --- | --- | --- | --- | --- |
|  |  |  |  |  | Min | Max | Mean |  | Min | Max | Mean |
| 1 | 394,267 | 394,642 | No | 375 | 109 | 716 | 403 | 0 | 0 | 0 | 0 |
| 2 | 397,053 | 398,215 | No | 1162 | 1023 | 1814 | 1218 | 44 | 39 | 72 | 46 |
| 3 | 402,360 | 402,496 | No | 136 | 79 | 193 | 130 | 7 | 4 | 10 | 7 |
| 4 | 415,337 | 415,688 | No | 351 | 181 | 3344 | 1206 | 23 | 12 | 245 | 80 |
| 5 | 416,971 | 417,370 | No | 399 | 42 | 423 | 392 | 16 | 3 | 17 | 16 |
| 6 | 420,823 | 420,823 | Yes | 0 | 0 | 221 | 92 | 1 | 1 | 25 | 11 |
| 7 | 424,161 | 424,507 | No | 346 | 345 | 865 | 473 | 7 | 7 | 12 | 8 |
| 8 | 429,247 | 429,487 | No | 240 | 181 | 305 | 236 | 12 | 10 | 14 | 11 |
| 9 | 436,182 | 436,284 | No | 102 | 102 | 269 | 171 | 7 | 7 | 17 | 11 |
| 10 | 467,404 | 468,468 | No | 1064 | 420 | 1156 | 688 | 26 | 6 | 26 | 14 |
| 11* | 474,175 | 474,849 | Yes | 674 | 201 | 1000 | 517 | 54 | 15 | 73 | 38 |
| 12 | 481,766 | 482,041 | Yes | 275 | 119 | 433 | 217 | 16 | 6 | 25 | 13 |
| 13 | 484,071 | 484,444 | No | 373 | 77 | 255 | 128 | 26 | 5 | 17 | 9 |
| 14 | 486,852 | 487,551 | Yes | 699 | 86 | 1313 | 295 | 34 | 7 | 72 | 17 |
| 15 | 492,113 | 492,414 | No | 301 | 207 | 493 | 250 | 10 | 5 | 14 | 7 |
| 16 | 495,383 | 495,935 | Yes | 552 | 167 | 1005 | 570 | 81 | 26 | 146 | 83 |
| 17 | 497,705 | 497,890 | Yes | 185 | 53 | 188 | 131 | 16 | 5 | 16 | 12 |
| 18 | 502,851 | 502,931 | Yes | 80 | 46 | 157 | 87 | 6 | 2 | 14 | 7 |
| 19 | 505,349 | 505,734 | No | 385 | 266 | 911 | 326 | 6 | 3 | 12 | 4 |
| 20 | 507,678 | 507,937 | Yes | 259 | 282 | 1618 | 542 | 25 | 27 | 184 | 60 |
| 21 | 512,320 | 512,467 | No | 147 | 3 | 150 | 148 | 3 | 0 | 3 | 3 |
| 22 | 515,765 | 515,975 | No | 210 | 156 | 294 | 213 | 0 | 0 | 1 | 0 |
| 23 | 528,198 | 528,391 | Yes | 193 | 192 | 241 | 214 | 15 | 15 | 21 | 19 |
| 24 | 532,188 | 532,527 | No | 339 | 219 | 341 | 325 | 3 | 1 | 3 | 3 |
| 25 | 536,408 | 537,076 | No | 668 | 451 | 744 | 644 | 22 | 15 | 23 | 21 |
| 26 | 542,226 | 542,281 | No | 55 | 29 | 557 | 244 | 6 | 4 | 52 | 23 |
| 27 | 543,248 | 543,292 | No | 44 | 3 | 343 | 43 | 1 | 0 | 9 | 1 |
| 28 | 548,865 | 549,215 | No | 350 | 202 | 646 | 398 | 19 | 10 | 26 | 18 |
| 29 | 552,154 | 553,838 | No | 1684 | 1198 | 3286 | 2987 | 1 | 1 | 17 | 2 |
| 30 | 554,781 | 555,055 | Yes | 274 | 53 | 1076 | 446 | 16 | 4 | 69 | 28 |
| 31 | 557,543 | 558,010 | No | 467 | 147 | 472 | 390 | 16 | 5 | 17 | 13 |
| 32 | 558,691 | 559,091 | No | 400 | 327 | 409 | 400 | 22 | 19 | 23 | 21 |
| 33 | 561,767 | 565,822 | Yes | 4055 | 1168 | 6003 | 2902 | 416 | 119 | 617 | 309 |
| 34 | 570,642 | 570,879 | No | 237 | 165 | 362 | 237 | 14 | 10 | 21 | 14 |
| 35 | 571,243 | 571,278 | No | 35 | 35 | 67 | 35 | 1 | 1 | 1 | 1 |
| 36 | 575,043 | 575,079 | No | 36 | 35 | 253 | 73 | 2 | 2 | 27 | 6 |
| 37 | 579,754 | 579,804 | No | 50 | 49 | 75 | 56 | 2 | 2 | 3 | 2 |
| 38 | 605,215 | 605,543 | No | 328 | 252 | 451 | 322 | 5 | 5 | 9 | 5 |
|  |  |  |  |  | Min | Max | Mean |  | Min | Max | Mean |
| 39 | 610,942 | 610,942 | No | 0 | 0 | 372 | 3 | 0 | 0 | 0 | 0 |
| 40 | 628,401 | 633,452 | No | 5051 | 2904 | 8041 | 4364 | 138 | 73 | 223 | 119 |
| 41 | 635,822 | 635,823 | No | 1 | 1 | 159 | 36 | 1 | 1 | 6 | 2 |
| 42 | 641,801 | 642,208 | No | 407 | 255 | 484 | 396 | 21 | 14 | 26 | 21 |
| 43 | 648,293 | 651,312 | No | 3019 | 2239 | 8952 | 4662 | 82 | 60 | 249 | 129 |
| 44 | 661,257 | 661,371 | No | 114 | 114 | 254 | 150 | 1 | 1 | 2 | 1 |
| 45<sup>†</sup> | 662,944 | 663,131 | Yes | 187 | 94 | 664 | 280 | 23 | 13 | 64 | 32 |
| 46 | 667,784 | 668,381 | Yes | 597 | 212 | 1172 | 412 | 56 | 21 | 112 | 38 |
| 47 | 675,125 | 675,145 | No | 20 | 6 | 54 | 33 | 0 | 0 | 0 | 0 |
| 48 | 675,455 | 675,486 | No | 31 | 31 | 79 | 45 | 0 | 0 | 2 | 1 |
| 49 | 676,820 | 677,020 | Yes | 200 | 151 | 206 | 195 | 35 | 26 | 37 | 34 |

All coordinates are based on HG38.
\* Extended position: Chr5:474,089-474,863
<sup>†</sup> Extended position: Chr5:662,841-663,226

### VNTR Associations with gene expression in CF nasal epithelia

Average lengths of VNTRs #44, 45 and 48 were associated with *TPPP* expression exceeding the multiple hypothesis testing corrected significance threshold [Figure 2]. Of these three VNTRs which were highly correlated [Table S4], only VNTR #45 overlaps a CpG island whereas both VNTR #44 and #48 include ≤2 CpGs. Both longer length of VNTR #45 (β (SE)=-0.002 (0.001), p=2.43×10^-4^) and higher CpG dinucleotide content (β (SE)=-0.025 (0.006), p=3.26×10^-4^) were associated with lower expression of *TPPP* explaining 5% of its variation [Table S5 & S6]. Longer length of VNTR #45 (β (SE) = 0.003 (0.001), p = 0.028) and its higher number of CpGs (β (SE) = 0.031 (0.013), p = 0.026) were also nominally associated with higher expression of *SLC9A3* explaining 8% of its variation. *TPPP* expression ranged from 1.9 to 64.2 TPM (median = 34.4); and *SLC9A3* expression ranged from 1.1 to 179.2 TPM (median = 14.2) [Table S7].

We used the average length of the two VNTR copies for each individual to test their association with gene expression. Using the shorter or longer length of the two copies instead of the average showed similar associations between VNTR #45 and *TPPP* and *SLC9A3* expression [Table S8].

VNTR #45 is located in the 3’ UTR of *TPPP* (oriented on the reverse strand). Its repeat unit is 31 nucleotides (CGGAATCACCCGATCACAGACGAGCGGTCAC) including 4 or 5 CpG copies depending upon SNP alleles within the repeat unit. Investigating the flanking sequence of the polymorphic portion of the VNTR (Chr5:662,944-663,131) revealed that it actually spans a region (Chr5:662,841-663,226) with 6 additional repeat units plus a 14 bp sequence similar to the first half of the repeat unit (CGGAATCACCCGAT), which is not variable and is present in all individuals [Figure 3A]. Participants had 9 to 27 (mean = 15) copies of the repeat unit on each haplotype. *TPPP* expression on average decreased by 1.4 TPM (8.4% of the standard deviation (SD)) per single repeat unit increase in the length of VNTR #45.

**Figure 3:**
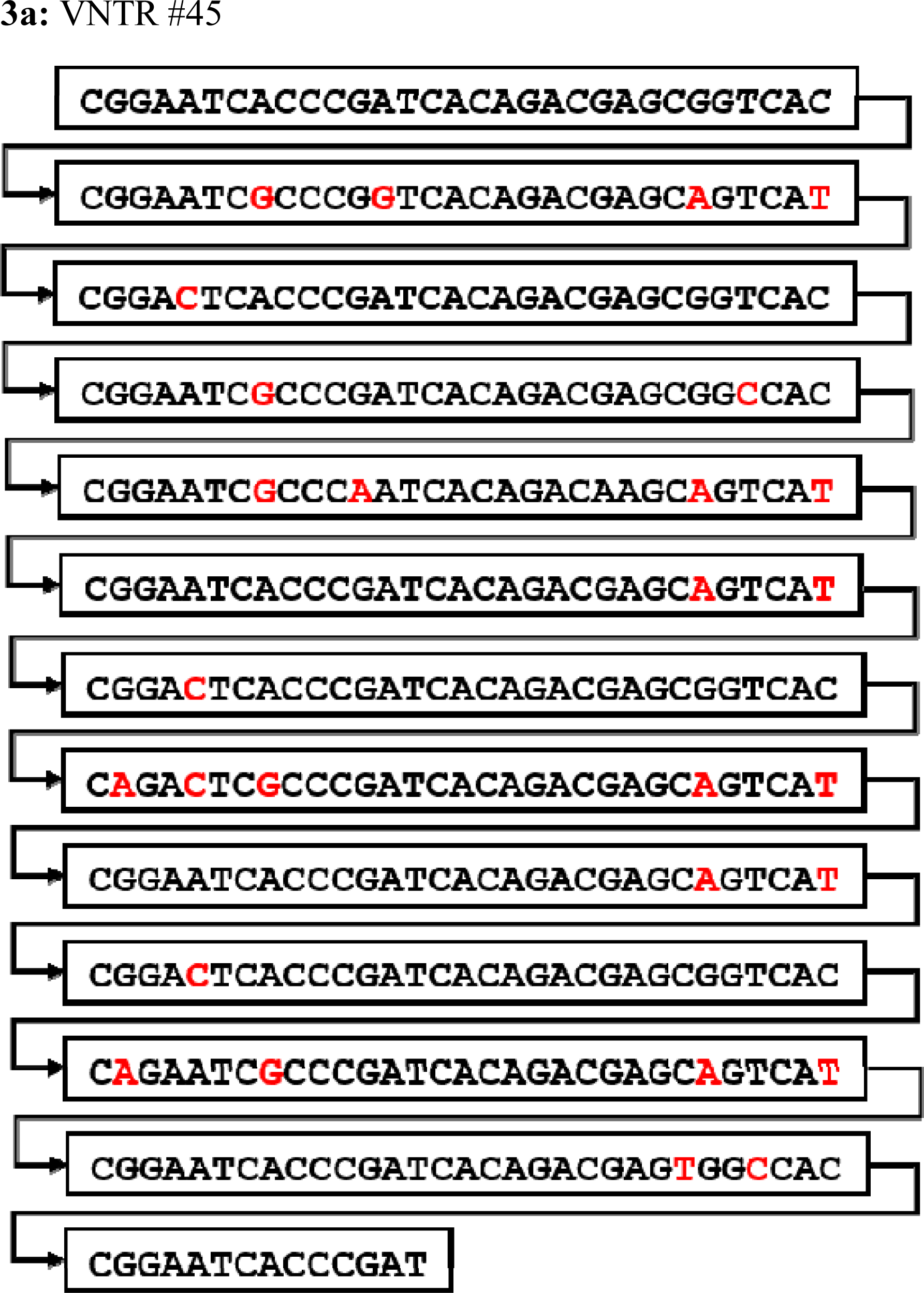
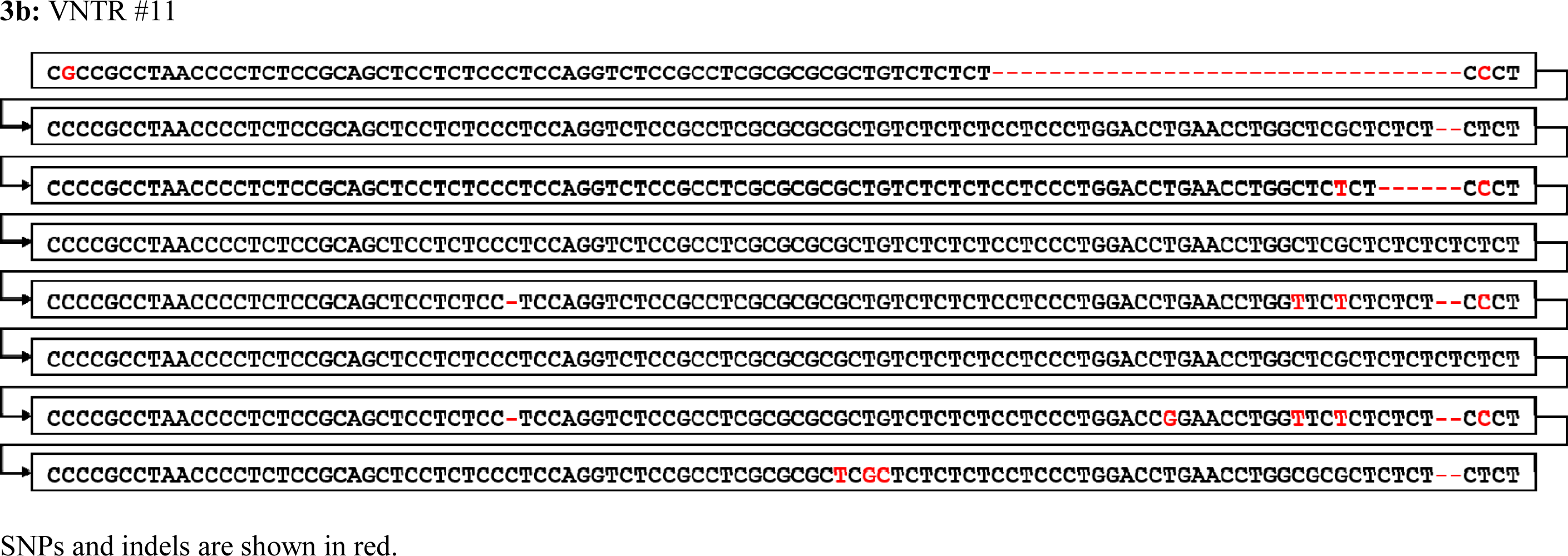
A depiction of VNTR #45 and #11 sequence in HG38 on the reverse strand

Given the distribution of lengths, VNTR #45 could be categorized into two groups: short ≤500 bp or ≤ 15 repeat units (frequency = 80%) and long > 500 bp or > 15 repeat units (frequency 20%). Longer length of the repeat is more frequently in *cis* with the minor allele (G) of rs57221529 (A>G, minor allele frequency (MAF) = 21%), the top SNP from the GWAS peak 5’ of *SLC9A3* (Fisher’s exact test p = 1.18×10^-15^) (r^2^ = 0.65, D’ = 0.83) [Table 3]. The G allele of rs57221529 was associated with reduced *TPPP* (β (SE) = -0.27 (0.09), p = 4.96×10^-3^) and increased *SLC9A3* expression (β (SE) = 0.36 (0.17), p = 3.37×10^-2^) in CF nasal epithelia explaining 2% and 3% of their variation, respectively, notably less than the proportions explained by VNTR #45 features.

Nine VNTRs all including CpGs (#10, 11, 13, 26, 36, 44, 45, 48, and 49) were nominally associated with *SLC9A3* expression. When these VNTRs were joint analyzed in a multivariable model, only VNTR #11 remained significant exceeding the multiple hypothesis testing corrected significance threshold (p <0.001). Both longer length of VNTR #11 (β (SE)=0.005 (0.001), p=6.85×10^-5^) and increased CpG dinucleotide content (β (SE)=0.058 (0.015), p=9.43×10^-4^) were associated with higher expression of *SLC9A3* explaining 12% of its variation [Table S9 & S10].

Using the shorter or longer length of the two VNTR copies for each individual instead of the average showed similar associations between VNTR #11 and *SLC9A3* expression [Table S8].

VNTR #11 is located in the last intron (intron 16) of *SLC9A3*. Its repeat unit is 97-103 nucleotides (CCCCGCCTAACCCCTCTCCGCAGCTCCTCTCCCTCCAGGTCTCCGCCTCGCGCGCGC TGTCTCTCT**CCTCCCTGGACCTGAACCTGGCTCGCTCTCTCT**CTCT) including 8-10 CpGs. The 31nucleotide segment demarcated in bold is missing completely in some repeat units, but when present includes varying numbers of CT dinucleotide repeats (2-8). Investigating the flanking sequence of the polymorphic portion of the VNTR (Chr5:474,175-474,849) revealed an additional repeat unit present in all individuals (Chr5:474,089-474,863) [Figure 3B]. Participants had 3 to 11 (mean = 6) copies of the repeat unit on each haplotype. *SLC9A3* expression on average increased by 10 TPM (28% of the SD) per single repeat unit increase in the length of VNTR #11.

Given the distribution of its length, VNTR #11 can be categorized into two sub-groups: short ≤ 550 bp or ≤ 5 repeat units (frequency = 21%) and long > 550 bp or > 5 repeat units (frequency 79%). The long lengths of the repeat are more frequently in *cis* with the major allele (A) of rs72711364 (A>G, MAF = 28%), the top SNP from the GWAS peak 3’ of *SLC9A3* (Fisher’s exact test p = 2.80×10^-12^) (r^2^ = 0.53, D’ = 0.88) [Table 3]. rs72711364 was not significantly associated with *SLC9A3* expression in CF nasal epithelia (β(SE) = 0.14 (0.15), p = 0.35) despite the strong association of VNTR #11 in this same sample.

Given that VNTRs #11 and #45 were tagged by the two top CF lung function GWAS associated variants and were independent of each other (Spearman correlation coefficient = 0.08, p = 0.40), we included both in a multivariable model to predict gene expression. Length and CpG number of both VNTR #11 and VNTR #45 remained significantly associated with *SLC9A3* expression, and the number of CpGs in the two VNTRs together explained 24% of the variation in *SLC9A3* expression [Table 2].

**Table 2:** Joint association of VNTR #11 and #45 (A: length, B: CpG number) with gene expression in CF nasal epithelia

| Gene | VNTR #11 |  |  | VNTR #45 |  |  | VNTR #11 & #45<br>R<sup>2</sup> |
| --- | --- | --- | --- | --- | --- | --- | --- |
|  | Beta | SE | P | Beta | SE | P |  |
| **AHRR** | -0.001 | 0.001 | 0.65 | -0.003 | 0.001 | 3.37E-2 | 0.06 |
| **EXOC3** | 0.003 | 0.001 | 1.22E-2 | 0.000 | 0.001 | 0.82 | 0.09 |
| **SLC9A3** | 0.004 | 0.001 | 5.15E-4 | 0.003 | 0.001 | 2.31E-3 | 0.22 |
| **CEP72** | 0.001 | 0.001 | 0.37 | 0.000 | 0.001 | 0.98 | 0.01 |
| **TPPP** | -0.001 | 0.001 | 0.08 | -0.002 | 0.001 | 8.95E-5 | 0.06 |

2b: CpG number
| Gene | VNTR #11 |  |  | VNTR #45 |  |  | VNTR #11 & #45<br>R<sup>2</sup> |
| --- | --- | --- | --- | --- | --- | --- | --- |
|  | Beta | SE | P | Beta | SE | P |  |
| **AHRR** | -0.009 | 0.016 | 0.60 | -0.031 | 0.013 | 2.36E-2 | 0.07 |
| **EXOC3** | 0.041 | 0.016 | 1.42E-2 | -0.002 | 0.013 | 0.872 | 0.09 |
| **SLC9A3** | 0.057 | 0.014 | 2.59E-4 | 0.040 | 0.011 | 1.09E-3 | 0.24 |
| **CEP72** | 0.013 | 0.014 | 0.35 | 0.001 | 0.001 | 0.95 | 0.01 |
| **TPPP** | -0.016 | 0.008 | 4.77E-2 | -0.027 | 0.006 | 7.96E-5 | 0.06 |

Association of the mean length and CpG number (of two haplotypes) of VNTRs with gene expression were tested using linear regression with both VNTRs, and sex, RIN, PC1-3, first 7 peer factors and *CD45* expression as covariates in the model.

**Table 3:**
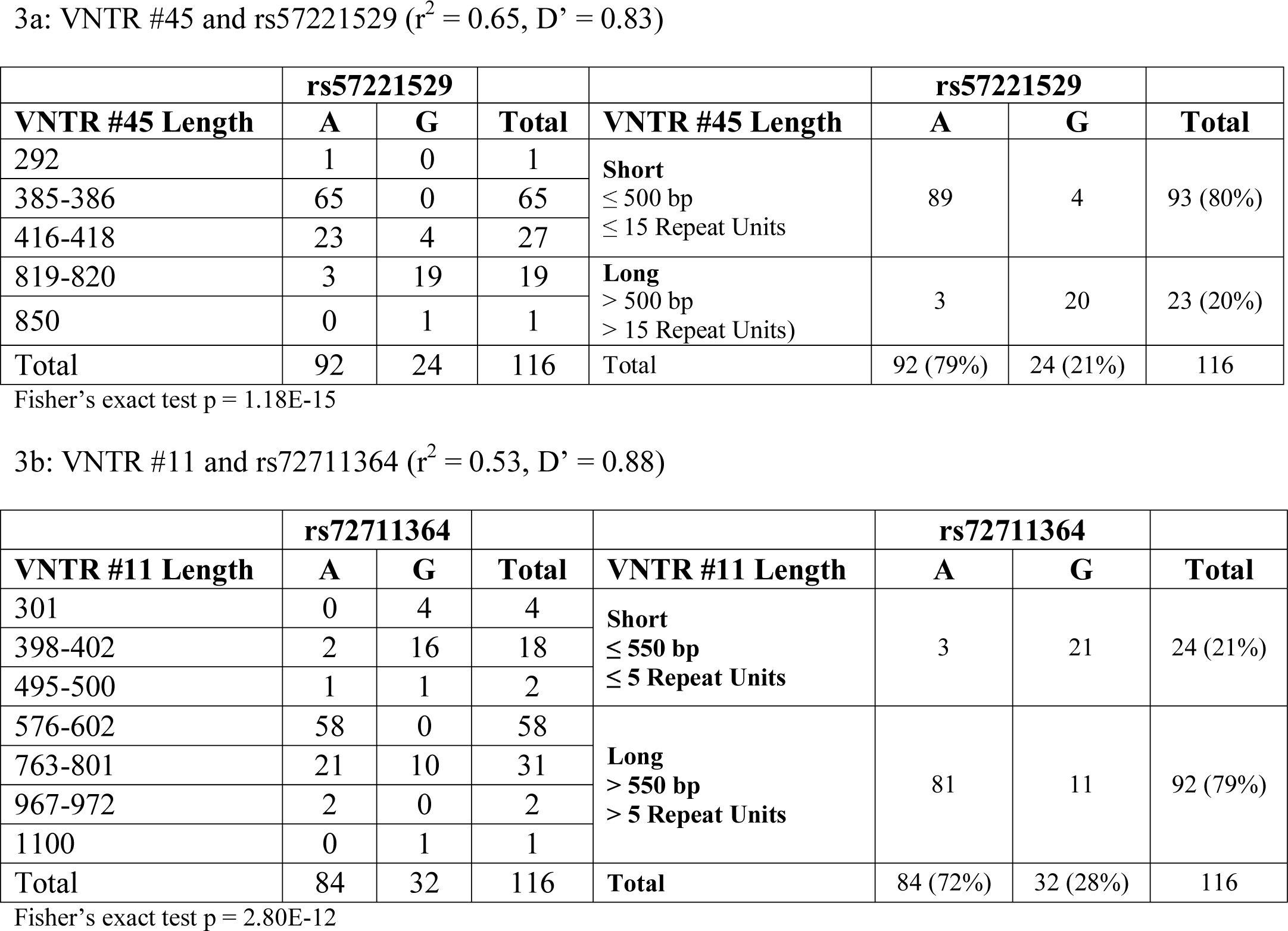
Presence of VNTR #45 and #11 with alleles of the top SNPs from the GWAS peaks 5’ and 3’ of *SLC9A3* (rs57221529 and rs72711364) on the same haplotype based on PacBio phased data

None of the 49 VNTRs were associated with *AHRR*, *EXOC3* and *CEP72* expression at the 0.001 significance threshold [Table S5 & S6].

### Estimating VNTR #11 and #45 lengths using 10XG short-read sequence

We then attempted to determine estimations of the VNTR #11 and #45 lengths from short-read sequence by dividing the number of reads aligned to the location of each VNTR (determined by PacBio) by the average sequencing depth. The estimated lengths of both VNTR #11 and #45 from the 10XG data were correlated with their corresponding lengths from PacBio (N = 52; Spearman correlation coefficients = 0.66 and 0.78, p = 9.38×10^-8^ and p = 1.43×10^-11^, respectively), providing “imputed” VNTR length calls from short-read sequence, even if not perfect estimates.

### Association of VNTR #45 and #11 with gene expression in GTEx

We estimated the lengths of both VNTR #45 and #11 in GTEx short-read sequence as described above and then assessed the association between these imputed VNTR lengths and gene expression across the 49 GTEx tissues, and in particular with lung expression for replication.

VNTR #45 was associated with *TPPP* and *SLC9A3* expression in multiple GTEx tissues [Table S11]. Longer imputed lengths of VNTR #45 were associated with higher *SLC9A3* expression in the GTEx lung tissue (β (SE) = 0.08 (0.03), p =1.09×10^-3^), explaining ∼1.2% of its variation whereas imputed lengths of VNTR #45 were not associated with *TPPP* expression in the GTEx lung tissue [Figure 4, 5]. Longer imputed VNTR #11 lengths were associated with lower expression of *SLC9A3* in most tissues in GTEx including the GTEx lung (β (SE) = -0.18 (0.02), p = 9.80×10^-13^), explaining 5% of its variation in this tissue [Figure 4, Table S12]. This is in contrast to the observation in CF nasal epithelia, where observed longer VNTR #11 lengths were significantly associated with higher *SLC9A3* expression. Imputed VNTR #11 and #45 lengths together explained 9% of *SLC9A3* expression variation in the non-CF GTEx lung samples.

**Figure 4:**
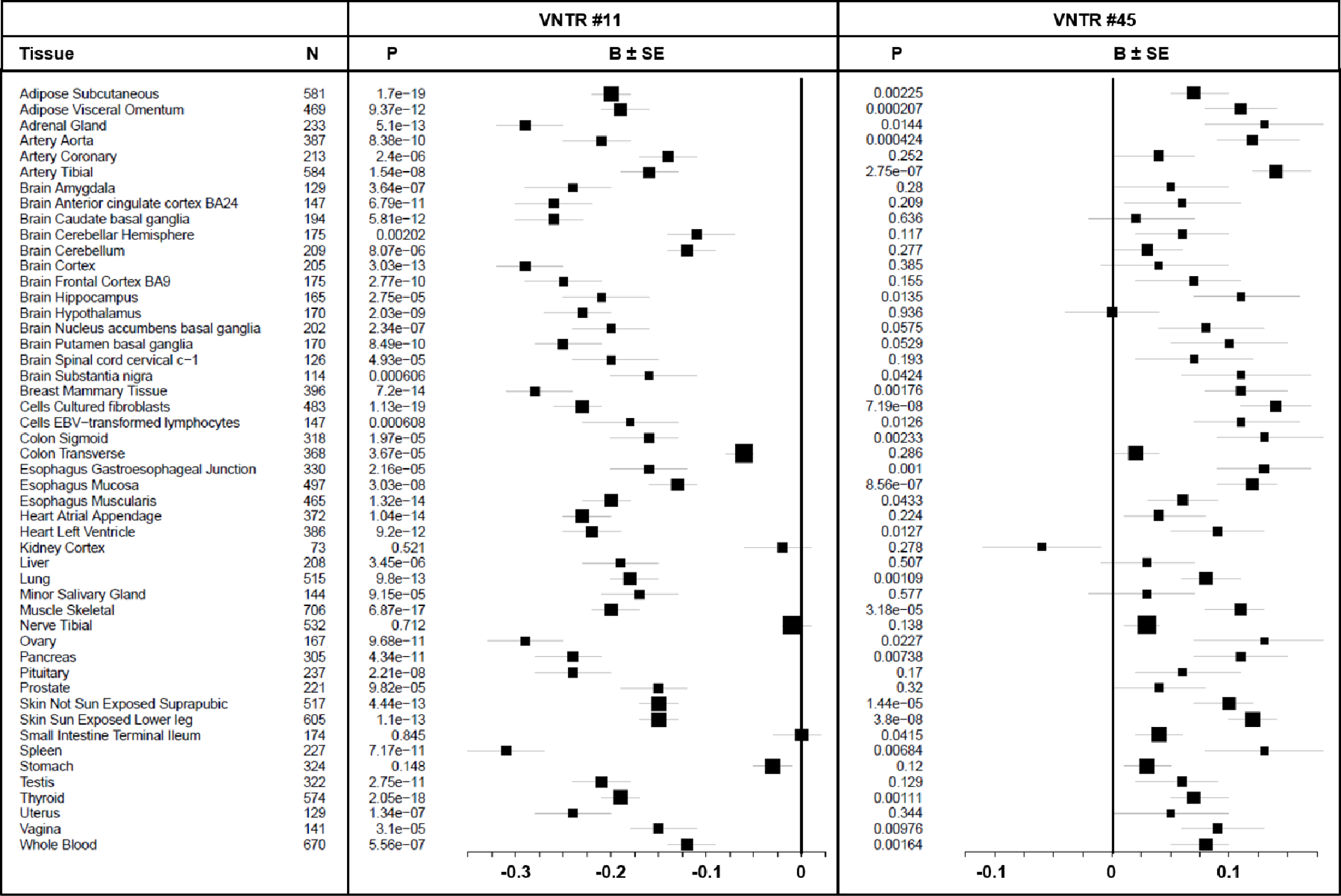
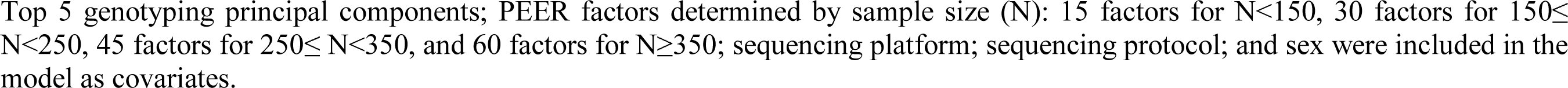
Association of VNTR #11 and #45 with *SLC9A3* expression in GTEx tissues

**Figure 5:**
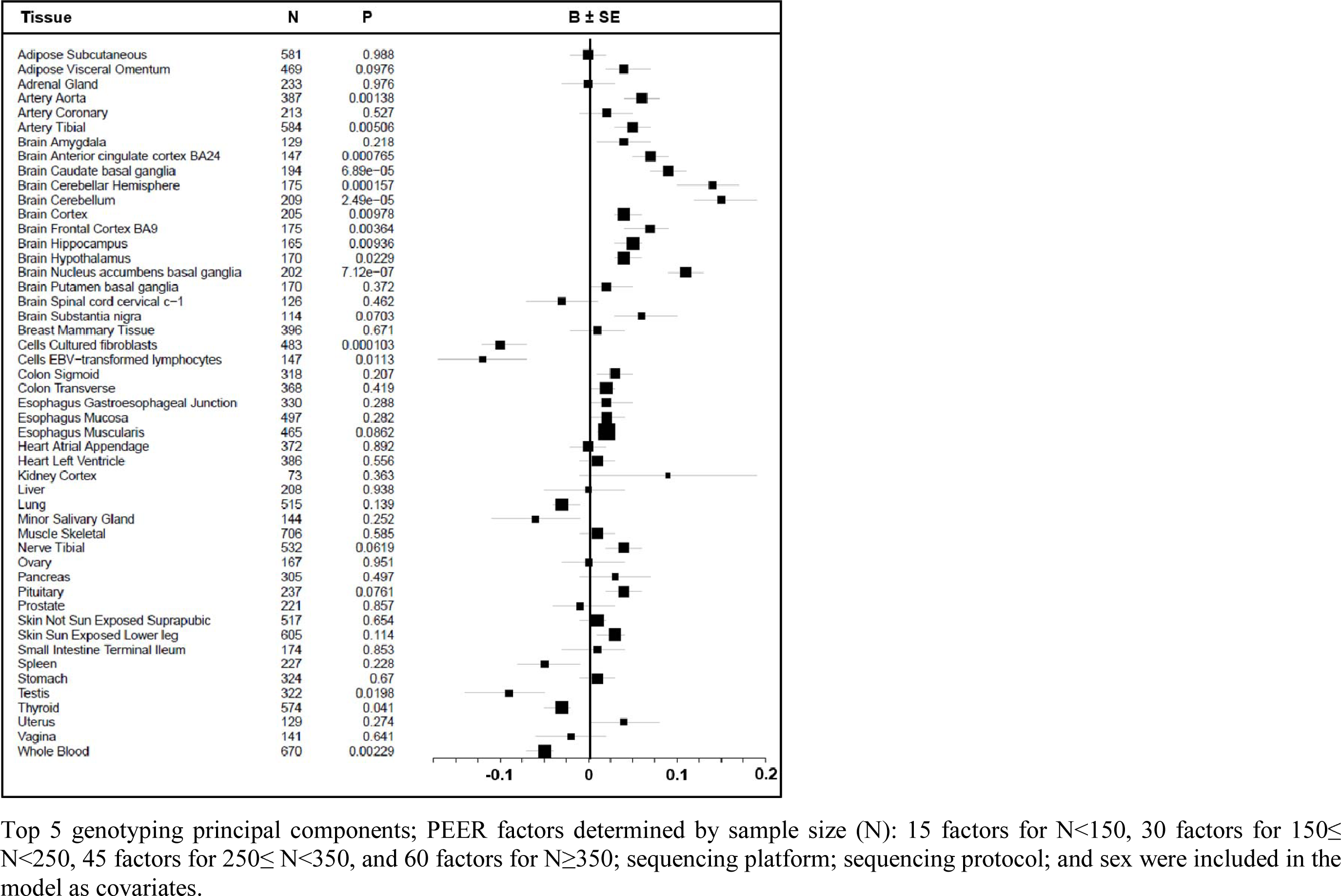
Association of VNTR #45 with *TPPP* expression in GTEx tissues

### *SLC9A3* expression in CF versus non-CF nasal epithelia

It was perplexing to see almost all of the GTEx tissues demonstrate VNTR#11 as a significant eVNTR with longer length resulting in less *SLC9A3* expression, while in CF nasal epithelia longer length resulted in significantly more *SLC9A3* expression. Since *SLC9A3* has previously been reported to associate with earlier acquisition of PsA in individuals with CF ^24; 48^, whereas this infection is not present in individuals without CF, we investigated whether *SLC9A3* expression differs in nasal epithelia between 6 non-CF individuals, 29 individuals with CF who also had an active PsA infection at the time of nasal brush and 39 individuals with CF without an active PsA infection. *SLC9A3* expression was significantly lower in CF patients with active PsA infections compared to non-CF individuals (β(SE) = -0.77 (0.33), p= 0.027), whereas gene expression in CF individuals without an active infection were similar to the non-CF sample (β (SE) = -0.07 (0.43), p = 0.86) [Figure 6].

**Figure 6:**
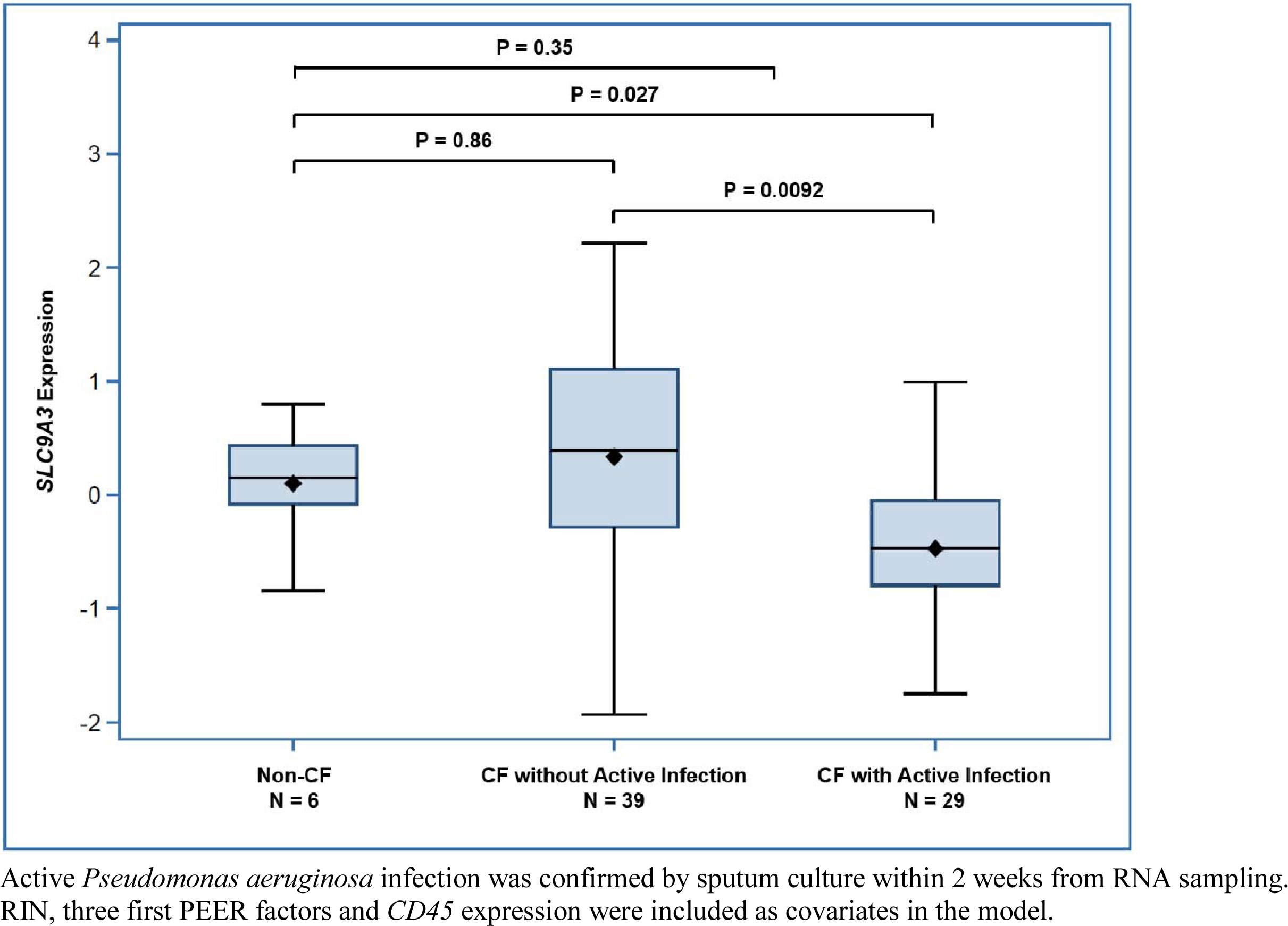
S*L*C9A3 expression in non-CF individuals vs. CF patients with or without active *Pseudomonas aeruginosa* infection

## DISCUSSION

By using mQTL and eQTM summary statistics from nasal epithelia in non-CF individuals, we demonstrated that a large proportion of SNPs associated with CF lung function at the genome-wide significance threshold are also associated with methylation levels at nearby CpGs that impact *SLC9A3* expression ^6; 35^. These SNPs are annotated to the two independent 5’ and 3’ CF lung function GWAS peaks at the *SLC9A3* locus, including the top GWAS associated SNPs, rs57221529 and rs7271136. These findings support the hypothesis that polymorphisms from both peaks impact lung function by altering methylation levels at CpGs that in turn influence *SLC9A3* expression. However, the GWAS and methylation studies used array-based technologies that analyzed SNPs and therefore do not provide the data necessary to determine the responsible causal variation given the complexity of tandem repeats at the locus ^36^.

A number of methods have been developed to genotype VNTRs genome-wide using next generation short and/or long-read sequencing. These methods are often computationally expensive and may not be very precise ^1; 49–52^. In contrast, here, we were investigating a targeted region. Therefore, we were able to define the exact boundaries of the VNTRs and their lengths on both haplotypes for each individual with CF by implementing a novel approach that uses multiple alignment of PacBio long-read reference aligned phased sequence. Although this approach may not be immediately feasible genome-wide, it provides high accuracy for better understanding of targeted regions within the genome.

We identified 49 common (frequency >2%) VNTRs at the chromosome 5 CF lung function GWAS locus, some spanning up to ≈9 kbp. This is reflective of a high density of VNTRs in the region as, on average, about 4-5 VNTRs are expected in a region of similar length ^5^. The majority of these VNTRs have CpGs within their repeat units and fourteen of them overlap CpG islands. This was expected given the CpG richness of the region, which includes a large number of CpG islands compared to the average on chromosome 5 or the whole genome ^33; 34^. In a subset of individuals with PacBio long-read sequence data, we demonstrated that two of these VNTRs overlapping CpG islands, VNTR #11 and #45, are tagged by the top associated SNPs from the 5’ and 3’ *SLC9A3* GWAS peaks and are independently associated with *SLC9A3* gene expression variation from RNA-seq of CF nasal epithelia.

We estimated VNTR lengths in short-read sequence by dividing the number of reads aligned to a corresponding VNTR location defined by the PacBio long-read data, by the average genome-wide sequencing depth. Read depth has been used before to predict the length of tandem repeats by tools such as CNVnator ^53^ and these predicted VNTR lengths have shown good accuracy ^5^. We demonstrated that our approach to estimate the lengths of both VNTR #11 and #45 have good accuracy, using individuals with CF who were sequenced with both short- and long-read technologies. We thus used this approach to estimate the length of these two VNTRs in the short-read sequence provided by GTEx and investigated their association with gene expression across all GTEx tissues. Both VNTR #45 and #11 were associated with *SLC9A3* gene expression in the lungs as well as in multiple GTEx tissues.

A longer length of VNTR #45, which is located in the 3’UTR of *TPPP*, was associated with lower expression of *TPPP* in CF nasal epithelia and higher expression of *SLC9A3*. VNTR #45 is tagged by rs57221529 (A>G), the top GWAS SNP from the peak 5’ of *SLC9A3*. The long length of the VNTR (> 500 bp) is more frequently on the same haplotype as the minor allele (G) of the SNP, which was also associated with lower *TPPP* and higher *SLC9A3* expression in CF nasal epithelia. However, association of the SNP with expression of both *TPPP* and *SLC9A3*, despite larger sample size (n = 68 in SNP analysis vs. 46 in VNTR association), explained a much lower proportion of variation in expression compared to the VNTR (3% for SNP vs. 22% for VNTR). These results suggest that the VNTR is more likely to be the variation affecting CF lung function though altering gene expression, than the GWAS SNP.

We observed similar relationships between VNTR #45, and both *TPPP* and *SLC9A3* expression in GTEx non-CF tissues. A longer length of VNTR #45 was associated with greater *TPPP* expression in a number of blood related GTEx tissues similar to CF nasal epithelia, and was associated with less *TPPP* expression in a number of GTEx brain tissues. It was not, however, associated with *TPPP* expression in the lung. A longer length of VNTR #45 was also associated with greater *SLC9A3* expression in many GTEx tissues including the lung which provided concordant evidence, in the same direction, as observed in the CF nasal epithelia.

Although VNTR #45 is associated with *TPPP* expression, this gene is mostly expressed in brain related tissues based on GTEx and there is little evidence connecting it with lung function or CF related phenotypes, whereas SLC9A3 is an epithelial ion exchanger and has been associated with many CF related phenotypes specially in the intestine ^26–29^.

A longer length of VNTR #11, which is located in the last intron of *SLC9A3*, was associated with greater *SLC9A3* expression in CF nasal epithelia. VNTR #11 is tagged by rs72711364 (A>G), the top SNP from the GWAS peak 3’of *SLC9A3* ^6^. The long length of the VNTR is more frequently on the same haplotype as the major allele (A) of rs72711364, although this LD is not complete. Interestingly, rs72711364 was not associated with *SLC9A3* expression in CF nasal epithelia despite a larger sample size than included in the VNTR expression analysis, suggesting that the VNTR is the variation affecting CF lung function through altering *SLC9A3* expression rather than the SNP.

VNTR #11 also showed association with *SLC9A3* expression in most GTEx tissues including lung, with p-values as small as 1×10^-19^. However, these associations demonstrated the opposite direction of effect from what was observed in the CF nasal epithelia. One obvious difference between the GTEx tissues and the CF nasal epithelia, is that the CF nasal epithelia were obtained from individuals with CF. The airway environment in subjects with CF is different from that of healthy individuals and provides ripe conditions for bacterial infections such as PsA. The CF lungs also undergo significant changes over time in terms of disease progression and microbial infections ^54^, which could impact S*LC9A3* expression and also interact with the VNTR. Indeed, some eQTLs are not evident in naïve cells and can only be identified in response to a stimulus, referred to as response eQTLs ^55–57^. Response eQTLs represent genetic variations that are modified by environmental factors due to gene-environment interactions. Stimulating cells by environmental factors such as pro/anti-inflammatory situations ^58^, bacterial ^56; 57^ and viral infection ^55^ have also been shown to change the direction of effect for some eQTLs akin to what we observed here for *SLC9A3* in nasal epithelia. In alignment with this hypothesis, we show that *SLC9A3* expression is significantly decreased in the nasal epithelia of individuals with CF who have active PsA infection versus healthy controls, even after adjusting for immune cell composition.

The length and CpG numbers of VNTR #11 and #45 together explain a substantial proportion (22% and 24%, respectively) of variation in *SLC9A3* expression in CF nasal epithelia. The explained variation in *SLC9A3* expression in the GTEx lung (9%) is still substantial but falls short of the estimate in the CF nasal epithelia. This is likely due to multiple factors including: CF vs. non-CF tissue ^54^; epithelial cells being just a small proportion of cells in the GTEx lung samples ^59^; and most notably that the VNTR lengths in the GTEx analysis, imputed from short-read sequence, are imperfect estimates of the actual lengths.

Recently Garg et al. ^5^ used the simple repeats track from UCSC to define the location of ∼ 90k VNTRs (repeat units >10 bp and total length >100 bp) across all autosomal chromosomes, merging overlapping VNTRs. Then, they used CNVnator ^53^ to estimate the relative diploid copy number of each autosomal VNTR region in GTEx and in the Pediatric Cardiac Genomic Consortium (PCGC) study, where both whole-genome short-read sequence and peripheral blood methylomes were available. These GTEx and PCGC resources were used to identify eVNTRs and VNTRs associated with CpG methylation levels (mVNTRs), respectively. VNTR #11 and #45 coincide with the location of two of the VNTRs reported in this study (chr5:474201-474983 and chr5:662835-663227). Consistent with our findings, the VNTR overlapping VNTR #11 was associated with *SLC9A3* expression in most GTEx tissues, and the VNTR overlapping VNTR #45 was associated with *TPPP* expression in a number of brain related tissues [Table S13]. Notably, the VNTR that coincides with VNTR #11 was associated with methylation at three CpGs: cg21743597 (Chr5: 475,090), cg21159260 (Chr5: 475,774) and cg17233264 (Chr5: 476,290); and the VNTR overlapping VNTR #45 was associated with methylation at two CpGs:cg19576778 (Chr5: 654,910) and cg16624210 (Chr5: 671,319) confirming that both VNTR #11 and VNTR #45 are mVNTRs [Table S14] ^5^. These CpGs are all located within ±50kb but outside the VNTR locations. Since methylation data is derived from the Illumina EPIC array rather than sequencing, it cannot provide any information about methylation at CpGs located within the VNTRs.

The repeat regions have low mappability when aligning ChIP-seq (chromatin immunoprecipitation followed by sequencing) short-reads ^60^, but VNTR #11 is in close proximity to a region 3’ of *SLC9A3* (Chr5:472,845-473,425) showing strong regulatory marks suggestive of a putative promoter in both Encyclopedia of DNA Elements (ENCODE) ^61^ and the Roadmap Epigenomics Project (REMC) ^62^. This region also shows strong bidirectional transcription, a marker of accessible chromatin that tends to be correlated with promoter/enhancer activity ^63^ in FANTOM5’s track, which maps the 5’ and 3’ end of transcripts via Cap Analysis of Gene Expression (CAGE) ^64^. ReMap ^65^ also shows a large number of transcription factors that have been detected via ChIP-seq in this region, further confirming DNA accessibility and regulation. Given the proximity of the VNTR to this regulatory region, it may affect its 3D conformation and result in altered regulation. Micro-HiC experiments ^66^ also suggests that this regulatory region physically interacts with the region where VNTR #45 is located [Figure 7].

**Figure 7:**
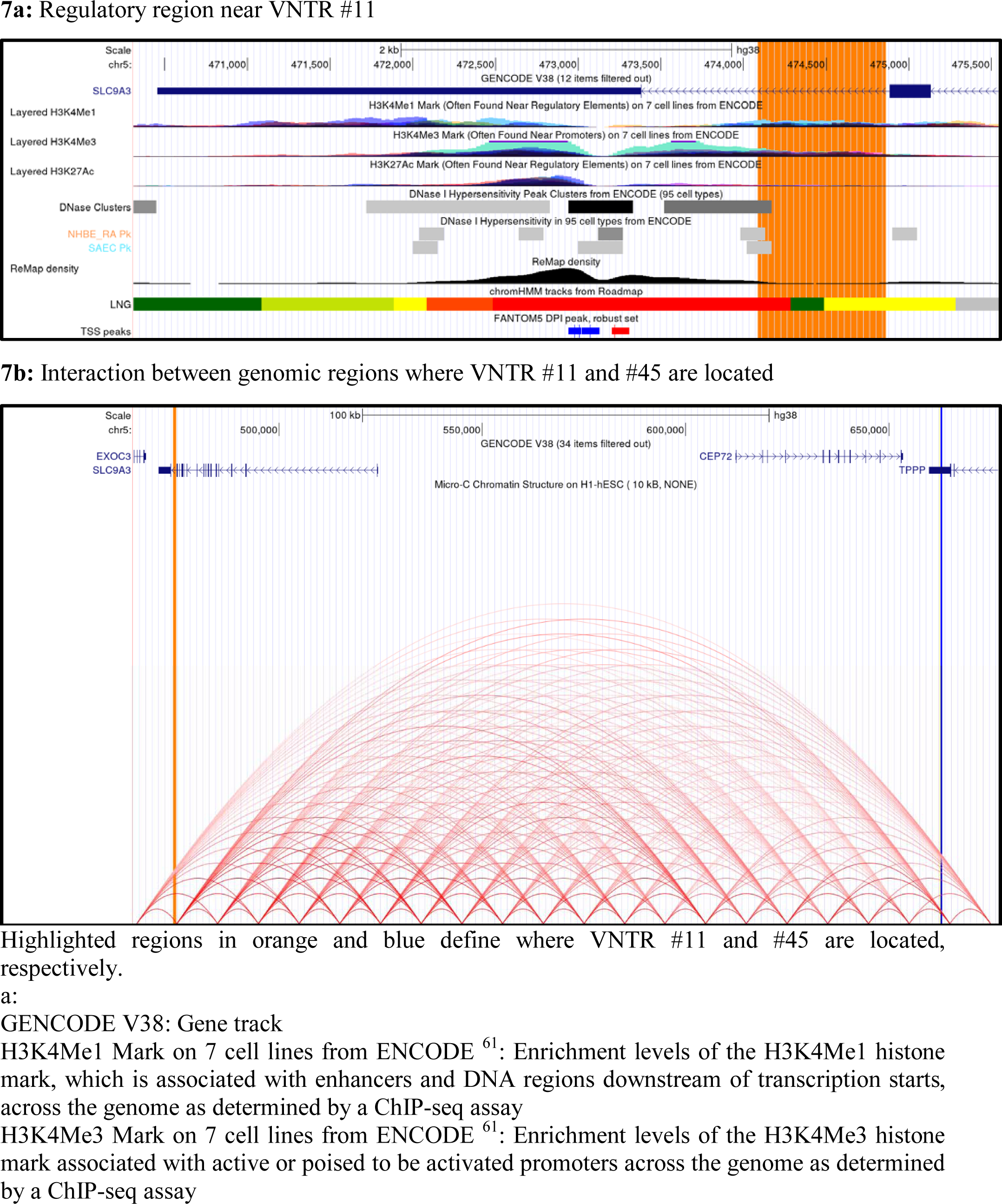
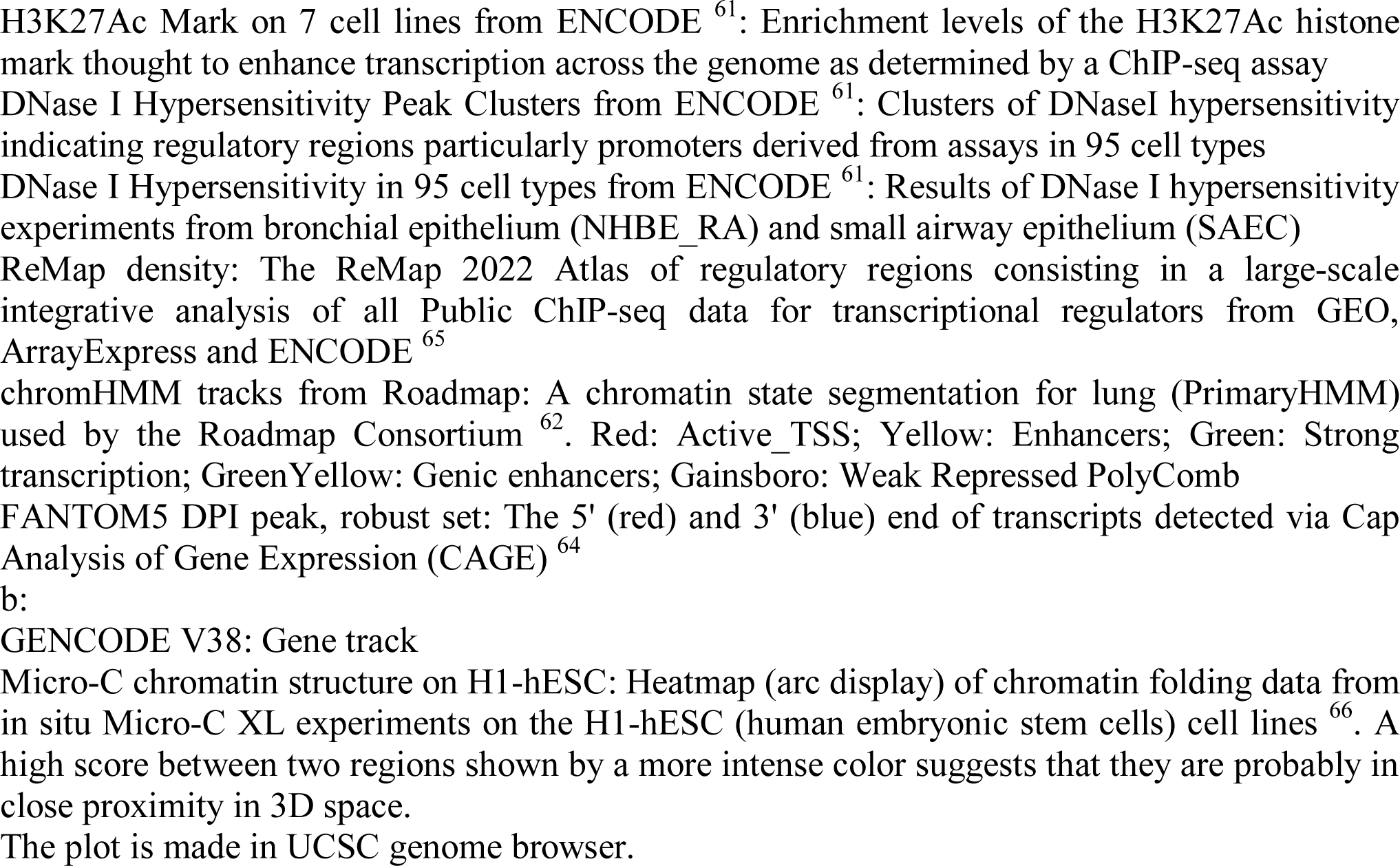
Evidence for a regulatory region near VNTR #11 and its interaction with the region where VNTR #45 is located

In conclusion, it is challenging to identify the causal variation responsible for GWAS signals in regions of genomic complexity, such as where there are a large number of VNTRs. Yet, our results demonstrate that VNTRs can lead to substantial gene expression variation and may explain appreciable missing heritability reported in complex traits. Long-read sequencing enables demarcation of the exact boundaries, lengths and repeat units of VNTRs, that are necessary to accurately estimate VNTR genotype from short-read sequence and fine map these loci. For VNTRs with CpGs within their repeat units, DNA methylation is a likely mechanism by which they affect gene expression; long-read technologies that simultaneously assess DNA methylation ^67^ will be an important technology to more directly address this moving forward.

## SUPPLEMENTAL INFORMATION DESCRIPTION

Supplementary file includes 14 tables and 2 figures.

Supplementary Excel file includes non-CF nasal epithelia mQTL and eQTM results.

## Supporting information

Nasal epithelia mQTL and eQTM data

## Data Availability

The RNAseq and whole genome sequence data from CF-affected individuals are available to researchers for academic, non-commercial research purposes through CFIT Program (https://lab.research.sickkids.ca/cfit/sequence-data-available/).

https://github.com/strug-hub/reference-polish

## ACKNOWLEDGEMENTS

Funding for this project was provided by Cystic Fibrosis Foundation STRUG17PO; Canadian Institutes of Health Research (FRN 167282); CF Canada (2626); the Program for Individualized CF Therapy (CFIT) funded by the SickKids Foundation and CF Canada; and Natural Sciences and Engineering Research Council of Canada (RGPIN: 2015-03742, 2013-250053). This work was also funded by the Government of Canada through Genome Canada (OGI-148) and supported by a grant from the Government of Ontario. The funders of the study play no role in study design, data collection and analysis, decision to publish or preparation of the manuscript.

The Genotype-Tissue Expression (GTEx) Project was supported by the Common Fund of the Office of the Director of the National Institutes of Health, and by NCI, NHGRI, NHLBI, NIDA, NIMH, and NINDS. The data used for the analyses described in this manuscript were obtained from dbGaP accession number phs000424.v8.p2 on Sep 2020.

## WEB RESOURCES

Cystic fibrosis mutation database: http://www.genet.sickkids.on.ca/StatisticsPage.html

CFTR2 website: https://cftr2.org/mutations_history

CF lung function meta-GWAS summary stats:

https://lab.research.sickkids.ca/strug/softwareandresources/

GTEx portal: https://gtexportal.org/home/datasets

The Human Protein Atlas: http://www.proteinatlas.org

UCSC Genome Browser: http://genome.ucsc.edu/index.html

pbmm2 v1.3.0: https://github.com/PacificBiosciences/pbmm2

Trim Galore v0.4.4: http://www.bioinformatics.babraham.ac.uk/projects/trim_galore

GCpp v1.9.0: https://github.com/PacificBiosciences/gcpp

Longshot v0.4.1: https://github.com/pjedge/longshot

MAFFT: https://mafft.cbrc.jp/alignment/software/

matSpD method: https://neurogenetics.qimrberghofer.edu.au/matSpD/

## DATA AND CODE AVAILABILITY

The scripts used to align PacBio reads to reference, phase and polish the sequences can be found at https://github.com/strug-hub/reference-polish.

