## Supplementary Tables and Figures for "A cystic fibrosis lung disease modifier locus harbors tandem repeats associated with gene expression"

### Supplementary Material

**Table S1:** Association of rs57221529 (A>G) with gene expressions in GTEx (V8)

| Gene | P-Value | NES | Tissue |
|---|---|---|---|
| AHRR | 3.50E-05 | -0.28 | Artery - Tibial |
| CEP72 | 3.50E-13 | -0.39 | Adipose - Subcutaneous |
| CEP72 | 2.00E-09 | -0.32 | Adipose - Visceral (Omentum) |
| CEP72 | 3.80E-06 | -0.44 | Adrenal Gland |
| CEP72 | 1.10E-07 | -0.4 | Artery - Aorta |
| CEP72 | 2.30E-11 | -0.32 | Artery - Tibial |
| CEP72 | 8.60E-13 | -0.31 | Breast - Mammary Tissue |
| CEP72 | 1.60E-08 | -0.39 | Colon - Sigmoid |
| CEP72 | 3.40E-06 | -0.28 | Colon - Transverse |
| CEP72 | 1.30E-08 | -0.39 | Esophagus - Gastroesophageal Junction |
| CEP72 | 5.60E-11 | -0.35 | Esophagus - Muscularis |
| CEP72 | 5.20E-06 | -0.23 | Lung |
| CEP72 | 6.60E-14 | -0.37 | Nerve - Tibial |
| CEP72 | 6.10E-08 | -0.43 | Prostate |
| CEP72 | 4.20E-05 | -0.2 | Skin - Not Sun Exposed (Suprapubic) |
| CEP72 | 6.80E-19 | -0.47 | Testis |
| CEP72 | 8.60E-14 | -0.31 | Thyroid |
| EXOC3 | 2.10E-06 | -0.2 | Lung |
| EXOC3 | 4.80E-05 | -0.24 | Spleen |
| EXOC3 | 1.80E-09 | -0.21 | Thyroid |
| EXOC3 | 3.40E-06 | -0.12 | Whole Blood |
| SLC9A3 | 8.90E-05 | 0.21 | Adipose - Subcutaneous |
| SLC9A3 | 7.80E-07 | 0.53 | Adrenal Gland |
| SLC9A3 | 1.60E-05 | 0.27 | Artery - Tibial |
| SLC9A3 | 6.70E-05 | 0.31 | Breast - Mammary Tissue |
| SLC9A3 | 2.20E-08 | 0.35 | Cells - Cultured fibroblasts |
| SLC9A3 | 5.50E-11 | 0.34 | Esophagus - Mucosa |
| SLC9A3 | 3.50E-07 | 0.42 | Heart - Left Ventricle |
| SLC9A3 | 1.10E-05 | 0.27 | Lung |
| SLC9A3 | 1.50E-07 | 0.3 | Muscle - Skeletal |
| SLC9A3 | 1.30E-05 | 0.54 | Ovary |
| SLC9A3 | 6.30E-05 | 0.36 | Pancreas |
| SLC9A3 | 3.20E-10 | 0.31 | Skin - Not Sun Exposed (Suprapubic) |
| SLC9A3 | 9.00E-17 | 0.4 | Skin - Sun Exposed (Lower leg) |
| SLC9A3 | 2.30E-07 | 0.27 | Thyroid |
| TPPP | 5.50E-06 | 0.22 | Artery - Aorta |
| TPPP | 1.20E-04 | 0.15 | Artery - Tibial |

| Gene | P-Value | NES | Tissue |
|---|---|---|---|
| TPPP | 8.00E-08 | 0.26 | Brain - Anterior cingulate cortex (BA24) |
| TPPP | 3.70E-08 | 0.27 | Brain - Caudate (basal ganglia) |
| TPPP | 2.30E-07 | 0.42 | Brain - Cerebellar Hemisphere |
| TPPP | 9.00E-14 | 0.54 | Brain - Cerebellum |
| TPPP | 1.80E-14 | 0.26 | Brain - Cortex |
| TPPP | 2.90E-05 | 0.21 | Brain - Frontal Cortex (BA9) |
| TPPP | 6.80E-09 | 0.26 | Brain - Nucleus accumbens (basal ganglia) |
| TPPP | 3.60E-07 | 0.25 | Brain - Putamen (basal ganglia) |
| TPPP | 6.20E-05 | 0.12 | Esophagus - Muscularis |
| TPPP | 3.80E-05 | -0.16 | Whole Blood |

**Table S2:** Association of rs72711364 (A>G) with gene expressions in GTEx (V8)

| **Gene** | **P-Value** | **NES** | **Tissue** |
|---|---|---|---|
| AHRR | 1.10E-07 | 0.19 | Cells - Cultured fibroblasts |
| AHRR | 6.50E-11 | 0.4 | Testis |
| CEP72 | 9.70E-06 | -0.22 | Testis |
| EXOC3 | 3.20E-12 | -0.22 | Adipose - Subcutaneous |
| EXOC3 | 2.60E-08 | -0.21 | Adipose - Visceral (Omentum) |
| EXOC3 | 3.50E-10 | -0.42 | Adrenal Gland |
| EXOC3 | 1.30E-13 | -0.34 | Artery - Aorta |
| EXOC3 | 1.10E-07 | -0.31 | Artery - Coronary |
| EXOC3 | 1.20E-17 | -0.24 | Artery - Tibial |
| EXOC3 | 6.80E-09 | -0.56 | Brain - Anterior cingulate cortex (BA24) |
| EXOC3 | 3.80E-07 | -0.4 | Brain - Cerebellar Hemisphere |
| EXOC3 | 5.70E-06 | -0.3 | Brain - Cerebellum |
| EXOC3 | 1.90E-07 | -0.41 | Brain - Frontal Cortex (BA9) |
| EXOC3 | 1.50E-07 | -0.43 | Brain - Hippocampus |
| EXOC3 | 4.40E-09 | -0.44 | Brain - Hypothalamus |
| EXOC3 | 3.20E-06 | -0.45 | Brain - Spinal cord (cervical c-1) |
| EXOC3 | 2.60E-08 | -0.55 | Brain - Substantia nigra |
| EXOC3 | 2.00E-10 | -0.24 | Breast - Mammary Tissue |
| EXOC3 | 1.60E-05 | -0.14 | Cells - Cultured fibroblasts |
| EXOC3 | 3.90E-16 | -0.39 | Colon - Sigmoid |
| EXOC3 | 6.30E-08 | -0.19 | Colon - Transverse |
| EXOC3 | 3.30E-11 | -0.27 | Esophagus - Gastroesophageal Junction |
| EXOC3 | 5.80E-18 | -0.3 | Esophagus - Mucosa |
| EXOC3 | 1.80E-23 | -0.38 | Esophagus - Muscularis |
| EXOC3 | 1.40E-10 | -0.3 | Heart - Atrial Appendage |
| EXOC3 | 1.40E-07 | -0.21 | Heart - Left Ventricle |
| EXOC3 | 3.20E-06 | -0.33 | Liver |
| EXOC3 | 9.80E-22 | -0.34 | Lung |
| EXOC3 | 4.00E-19 | -0.24 | Muscle - Skeletal |
| EXOC3 | 8.30E-19 | -0.31 | Pancreas |
| EXOC3 | 3.20E-06 | -0.31 | Prostate |
| EXOC3 | 6.00E-19 | -0.42 | Spleen |
| EXOC3 | 3.30E-13 | -0.31 | Stomach |
| EXOC3 | 1.50E-12 | -0.4 | Testis |
| EXOC3 | 1.30E-23 | -0.3 | Thyroid |
| EXOC3 | 1.50E-44 | -0.29 | Whole Blood |
| SLC9A3 | 2.60E-08 | 0.38 | Breast - Mammary Tissue |
| SLC9A3 | 9.20E-10 | 0.33 | Cells - Cultured fibroblasts |
| SLC9A3 | 1.10E-04 | 0.18 | Esophagus - Mucosa |
| SLC9A3 | 1.50E-05 | 0.25 | Esophagus - Muscularis |

| **Gene** | **P-Value** | **NES** | **Tissue** |
|---|---|---|---|
| SLC9A3 | 3.30E-07 | 0.35 | Heart - Atrial Appendage |
| SLC9A3 | 8.90E-13 | 0.52 | Heart - Left Ventricle |
| SLC9A3 | 1.40E-15 | 0.41 | Muscle - Skeletal |
| SLC9A3 | 6.20E-08 | 0.24 | Skin - Not Sun Exposed (Suprapubic) |
| SLC9A3 | 3.70E-15 | 0.34 | Skin - Sun Exposed (Lower leg) |
| SLC9A3 | 3.60E-06 | 0.43 | Spleen |
| SLC9A3 | 1.70E-08 | 0.26 | Thyroid |
| SLC9A3 | 2.40E-06 | 0.23 | Whole Blood |

**Table S3:** Correlation between the length and number of CpGs in each VNTR (N = 58)

| | Spearman Correlation | |
|---|---|---|
| **VNTR** | **Coefficient** | **P** |
| 1 | NA | NA |
| 2 | 0.77 | 1.83E-24 |
| 3 | 0.90 | 2.14E-42 |
| 4 | 0.99 | 6.50E-107 |
| 5 | 0.83 | 2.42E-31 |
| 6 | 1.00 | 1.43E-125 |
| 7 | 0.80 | 1.08E-26 |
| 8 | 0.49 | 3.11E-08 |
| 9 | 0.67 | 1.65E-16 |
| 10 | 0.98 | 1.41E-80 |
| 11 | 0.94 | 4.33E-57 |
| 12 | 0.87 | 1.64E-37 |
| 13 | 0.98 | 2.39E-78 |
| 14 | 0.99 | 1.10E-98 |
| 15 | 0.92 | 4.14E-49 |
| 16 | 0.99 | 2.51E-101 |
| 17 | 0.97 | 3.09E-68 |
| 18 | 0.94 | 4.88E-56 |
| 19 | 0.77 | 3.69E-24 |
| 20 | 0.87 | 4.54E-37 |
| 21 | 0.44 | 6.06E-07 |
| 22 | -0.55 | 2.42E-10 |
| 23 | 0.83 | 9.35E-31 |
| 24 | 0.92 | 1.69E-47 |
| 25 | 0.75 | 8.33E-22 |
| 26 | 0.98 | 1.31E-84 |
| 27 | 0.81 | 1.51E-28 |
| 28 | 0.24 | 8.87E-03 |
| 29 | -0.36 | 7.38E-05 |
| 30 | 0.84 | 5.26E-32 |
| 31 | 0.95 | 4.56E-59 |
| 32 | -0.17 | 7.46E-02 |
| 33 | 1.00 | 9.36E-118 |
| 34 | 0.42 | 3.08E-06 |
| 35 | NA | NA |
| 36 | 0.94 | 6.05E-57 |
| 37 | 0.98 | 1.74E-78 |
| 38 | 0.48 | 4.26E-08 |
| 39 | NA | NA |
| 40 | 0.97 | 9.63E-76 |

| | Spearman Correlation | |
|---|---|---|
| VNTR | Coefficient | P |
| 41 | 0.98 | 7.48E-88 |
| 42 | 0.96 | 3.80E-67 |
| 43 | 0.98 | 1.98E-82 |
| 44 | 0.76 | 2.01E-23 |
| 45 | 1.00 | 2.26E-127 |
| 46 | 0.96 | 8.83E-68 |
| 47 | NA | NA |
| 48 | 0.86 | 5.85E-35 |
| 49 | 0.63 | 3.75E-14 |

**Table S4:** Correlation between VNTR lengths (N = 58)

| | | | | | | | | | | | | | | | | | | | | | | | | | | | | | | | | | | | | | | | | | | | | | | | | |
|---|---|---|---|---|---|---|---|---|---|---|---|---|---|---|---|---|---|---|---|---|---|---|---|---|---|---|---|---|---|---|---|---|---|---|---|---|---|---|---|---|---|---|---|---|---|---|---|---|
| 1 | | | | | | | | | | | | | | | | | | | | | | | | | | | | | | | | | | | | | | | | | | | | | | | | |
| 0.42 | 2 | | | | | | | | | | | | | | | | | | | | | | | | | | | | | | | | | | | | | | | | | | | | | | | |
| 0.02 | 0.08 | 3 | | | | | | | | | | | | | | | | | | | | | | | | | | | | | | | | | | | | | | | | | | | | | | |
| 0.15 | 0.03 | 0.04 | 4 | | | | | | | | | | | | | | | | | | | | | | | | | | | | | | | | | | | | | | | | | | | | | |
| 0.18 | 0.00 | 0.11 | 0.16 | 5 | | | | | | | | | | | | | | | | | | | | | | | | | | | | | | | | | | | | | | | | | | | | |
| 0.00 | 0.19 | 0.29 | 0.13 | 0.07 | 6 | | | | | | | | | | | | | | | | | | | | | | | | | | | | | | | | | | | | | | | | | | | |
| 0.03 | 0.07 | 0.04 | 0.22 | 0.03 | 0.53 | 7 | | | | | | | | | | | | | | | | | | | | | | | | | | | | | | | | | | | | | | | | | | |
| 0.12 | 0.08 | 0.06 | 0.10 | 0.05 | 0.16 | 0.35 | 8 | | | | | | | | | | | | | | | | | | | | | | | | | | | | | | | | | | | | | | | | | |
| 0.21 | 0.20 | 0.03 | 0.10 | 0.14 | 0.29 | 0.10 | 0.41 | 9 | | | | | | | | | | | | | | | | | | | | | | | | | | | | | | | | | | | | | | | | |
| 0.04 | 0.23 | 0.04 | 0.15 | 0.01 | 0.68 | 0.37 | 0.04 | 0.50 | 10 | | | | | | | | | | | | | | | | | | | | | | | | | | | | | | | | | | | | | | | |
| 0.06 | 0.02 | 0.17 | 0.38 | 0.08 | 0.06 | 0.16 | 0.12 | 0.11 | 0.54 | 11 | | | | | | | | | | | | | | | | | | | | | | | | | | | | | | | | | | | | | | |
| 0.05 | 0.07 | 0.09 | 0.06 | 0.08 | 0.39 | 0.56 | 0.23 | 0.04 | 0.54 | 0.36 | 12 | | | | | | | | | | | | | | | | | | | | | | | | | | | | | | | | | | | | | |
| 0.11 | 0.09 | 0.13 | 0.44 | 0.33 | 0.08 | 0.17 | 0.16 | 0.45 | 0.26 | 0.15 | 0.36 | 13 | | | | | | | | | | | | | | | | | | | | | | | | | | | | | | | | | | | | |
| 0.00 | 0.08 | 0.08 | 0.09 | 0.02 | 0.57 | 0.63 | 0.21 | 0.07 | 0.60 | 0.40 | 0.58 | 0.10 | 14 | | | | | | | | | | | | | | | | | | | | | | | | | | | | | | | | | | | |
| 0.03 | 0.05 | 0.07 | 0.15 | 0.30 | 0.04 | 0.04 | 0.22 | 0.40 | 0.19 | 0.11 | 0.19 | 0.56 | 0.05 | 15 | | | | | | | | | | | | | | | | | | | | | | | | | | | | | | | | | | |
| 0.12 | 0.07 | 0.15 | 0.48 | 0.37 | 0.15 | 0.27 | 0.16 | 0.34 | 0.23 | 0.01 | 0.23 | 0.58 | 0.20 | 0.54 | 16 | | | | | | | | | | | | | | | | | | | | | | | | | | | | | | | | | |
| 0.05 | 0.14 | 0.18 | 0.12 | 0.11 | 0.46 | 0.38 | 0.00 | 0.27 | 0.41 | 0.12 | 0.13 | 0.11 | 0.47 | 0.09 | 0.25 | 17 | | | | | | | | | | | | | | | | | | | | | | | | | | | | | | | | |
| 0.10 | 0.01 | 0.17 | 0.37 | 0.05 | 0.32 | 0.30 | 0.00 | 0.18 | 0.09 | 0.11 | 0.03 | 0.36 | 0.01 | 0.31 | 0.22 | 0.39 | 18 | | | | | | | | | | | | | | | | | | | | | | | | | | | | | | | |
| 0.04 | 0.07 | 0.22 | 0.02 | 0.04 | 0.12 | 0.10 | 0.16 | 0.22 | 0.16 | 0.17 | 0.00 | 0.18 | 0.11 | 0.14 | 0.03 | 0.15 | 0.05 | 19 | | | | | | | | | | | | | | | | | | | | | | | | | | | | | | |
| 0.04 | 0.01 | 0.03 | 0.23 | 0.03 | 0.09 | 0.10 | 0.04 | 0.14 | 0.14 | 0.10 | 0.12 | 0.07 | 0.27 | 0.06 | 0.11 | 0.43 | 0.19 | 0.21 | 20 | | | | | | | | | | | | | | | | | | | | | | | | | | | | | |
| 0.08 | 0.04 | 0.09 | 0.16 | 0.03 | 0.04 | 0.10 | 0.05 | 0.00 | 0.11 | 0.13 | 0.11 | 0.26 | 0.26 | 0.06 | 0.01 | 0.07 | 0.30 | 0.21 | 0.36 | 21 | | | | | | | | | | | | | | | | | | | | | | | | | | | | |
| 0.01 | 0.09 | 0.27 | 0.14 | 0.00 | 0.21 | 0.04 | 0.08 | 0.01 | 0.05 | 0.10 | 0.13 | 0.08 | 0.15 | 0.13 | 0.08 | 0.07 | 0.21 | 0.10 | 0.06 | 0.05 | 22 | | | | | | | | | | | | | | | | | | | | | | | | | | | |
| 0.20 | 0.03 | 0.05 | 0.03 | 0.01 | 0.09 | 0.09 | 0.02 | 0.13 | 0.08 | 0.12 | 0.04 | 0.14 | 0.14 | 0.02 | 0.12 | 0.05 | 0.03 | 0.09 | 0.06 | 0.09 | 0.32 | 23 | | | | | | | | | | | | | | | | | | | | | | | | | | |
| 0.16 | 0.11 | 0.04 | 0.38 | 0.06 | 0.26 | 0.24 | 0.01 | 0.19 | 0.22 | 0.02 | 0.04 | 0.33 | 0.03 | 0.14 | 0.34 | 0.41 | 0.48 | 0.00 | 0.31 | 0.19 | 0.34 | 0.20 | 24 | | | | | | | | | | | | | | | | | | | | | | | | | |
| 0.00 | 0.04 | 0.09 | 0.25 | 0.12 | 0.16 | 0.11 | 0.01 | 0.16 | 0.09 | 0.00 | 0.12 | 0.21 | 0.07 | 0.21 | 0.23 | 0.22 | 0.40 | 0.02 | 0.22 | 0.10 | 0.51 | 0.24 | 0.71 | 25 | | | | | | | | | | | | | | | | | | | | | | | | |
| 0.21 | 0.23 | 0.06 | 0.10 | 0.01 | 0.12 | 0.01 | 0.16 | 0.29 | 0.15 | 0.08 | 0.13 | 0.19 | 0.19 | 0.16 | 0.05 | 0.26 | 0.45 | 0.02 | 0.15 | 0.29 | 0.08 | 0.32 | 0.46 | 0.36 | 26 | | | | | | | | | | | | | | | | | | | | | | | |
| 0.14 | 0.20 | 0.25 | 0.34 | 0.04 | 0.35 | 0.06 | 0.06 | 0.22 | 0.15 | 0.07 | 0.18 | 0.16 | 0.05 | 0.24 | 0.18 | 0.33 | 0.50 | 0.15 | 0.32 | 0.04 | 0.45 | 0.15 | 0.64 | 0.58 | 0.51 | 27 | | | | | | | | | | | | | | | | | | | | | | |
| 0.07 | 0.02 | 0.20 | 0.18 | 0.13 | 0.29 | 0.03 | 0.03 | 0.08 | 0.05 | 0.21 | 0.03 | 0.00 | 0.07 | 0.08 | 0.09 | 0.32 | 0.27 | 0.08 | 0.24 | 0.17 | 0.27 | 0.06 | 0.32 | 0.30 | 0.15 | 0.43 | 28 | | | | | | | | | | | | | | | | | | | | | |
| 0.03 | 0.01 | 0.05 | 0.10 | 0.17 | 0.21 | 0.15 | 0.18 | 0.14 | 0.11 | 0.06 | 0.04 | 0.05 | 0.17 | 0.22 | 0.25 | 0.16 | 0.20 | 0.05 | 0.21 | 0.20 | 0.24 | 0.41 | 0.28 | 0.45 | 0.08 | 0.38 | 0.33 | 29 | | | | | | | | | | | | | | | | | | | | |
| 0.17 | 0.25 | 0.07 | 0.16 | 0.04 | 0.19 | 0.01 | 0.03 | 0.16 | 0.10 | 0.12 | 0.06 | 0.06 | 0.05 | 0.20 | 0.09 | 0.26 | 0.36 | 0.04 | 0.33 | 0.09 | 0.22 | 0.05 | 0.51 | 0.52 | 0.34 | 0.57 | 0.67 | 0.41 | 30 | | | | | | | | | | | | | | | | | | | |
| 0.09 | 0.07 | 0.09 | 0.27 | 0.17 | 0.26 | 0.13 | 0.09 | 0.15 | 0.11 | 0.14 | 0.05 | 0.10 | 0.07 | 0.22 | 0.24 | 0.38 | 0.36 | 0.04 | 0.33 | 0.10 | 0.26 | 0.11 | 0.55 | 0.54 | 0.28 | 0.57 | 0.79 | 0.45 | 0.75 | 31 | | | | | | | | | | | | | | | | | | |
| 0.13 | 0.06 | 0.04 | 0.12 | 0.05 | 0.13 | 0.03 | 0.02 | 0.09 | 0.00 | 0.21 | 0.03 | 0.11 | 0.04 | 0.02 | 0.11 | 0.08 | 0.13 | 0.08 | 0.08 | 0.16 | 0.01 | 0.00 | 0.03 | 0.01 | 0.07 | 0.04 | 0.24 | 0.01 | 0.03 | 0.13 | 32 | | | | | | | | | | | | | | | | | |
| 0.08 | 0.01 | 0.11 | 0.33 | 0.06 | 0.19 | 0.12 | 0.03 | 0.12 | 0.03 | 0.16 | 0.04 | 0.15 | 0.02 | 0.29 | 0.27 | 0.28 | 0.37 | 0.21 | 0.30 | 0.01 | 0.30 | 0.24 | 0.38 | 0.32 | 0.03 | 0.47 | 0.57 | 0.46 | 0.52 | 0.62 | 0.20 | 33 | | | | | | | | | | | | | | | | |
| 0.14 | 0.11 | 0.05 | 0.04 | 0.00 | 0.12 | 0.01 | 0.03 | 0.02 | 0.05 | 0.02 | 0.11 | 0.08 | 0.04 | 0.00 | 0.01 | 0.03 | 0.18 | 0.11 | 0.02 | 0.15 | 0.05 | 0.06 | 0.11 | 0.03 | 0.17 | 0.15 | 0.03 | 0.02 | 0.05 | 0.03 | 0.15 | 0.00 | 34 | | | | | | | | | | | | | | | |
| 0.17 | 0.10 | 0.14 | 0.08 | 0.02 | 0.01 | 0.08 | 0.02 | 0.07 | 0.02 | 0.09 | 0.03 | 0.04 | 0.06 | 0.06 | 0.06 | 0.05 | 0.05 | 0.13 | 0.07 | 0.20 | 0.11 | 0.15 | 0.05 | 0.04 | 0.17 | 0.04 | 0.06 | 0.02 | 0.01 | 0.04 | 0.06 | 0.09 | 0.02 | 35 | | | | | | | | | | | | | | |
| 0.10 | 0.15 | 0.09 | 0.26 | 0.12 | 0.23 | 0.09 | 0.08 | 0.19 | 0.07 | 0.16 | 0.23 | 0.13 | 0.13 | 0.24 | 0.25 | 0.23 | 0.52 | 0.01 | 0.22 | 0.15 | 0.38 | 0.17 | 0.67 | 0.72 | 0.51 | 0.73 | 0.40 | 0.42 | 0.62 | 0.63 | 0.12 | 0.47 | 0.21 | 0.06 | 36 | | | | | | | | | | | | | |
| 0.09 | 0.15 | 0.19 | 0.32 | 0.08 | 0.28 | 0.06 | 0.10 | 0.20 | 0.07 | 0.22 | 0.09 | 0.09 | 0.08 | 0.18 | 0.24 | 0.35 | 0.43 | 0.07 | 0.34 | 0.07 | 0.28 | 0.05 | 0.49 | 0.39 | 0.37 | 0.58 | 0.73 | 0.28 | 0.68 | 0.73 | 0.17 | 0.66 | 0.11 | 0.08 | 0.70 | 37 | | | | | | | | | | | | |
| 0.10 | 0.05 | 0.02 | 0.07 | 0.04 | 0.15 | 0.15 | 0.08 | 0.02 | 0.09 | 0.07 | 0.04 | 0.09 | 0.20 | 0.21 | 0.13 | 0.20 | 0.13 | 0.06 | 0.11 | 0.10 | 0.04 | 0.11 | 0.09 | 0.21 | 0.11 | 0.05 | 0.23 | 0.18 | 0.10 | 0.23 | 0.04 | 0.42 | 0.15 | 0.02 | 0.05 | 0.27 | 38 | | | | | | | | | | | |
| 0.08 | 0.13 | 0.04 | 0.18 | 0.17 | 0.12 | 0.09 | 0.01 | 0.01 | 0.12 | 0.09 | 0.10 | 0.19 | 0.20 | 0.15 | 0.00 | 0.19 | 0.15 | 0.01 | 0.08 | 0.18 | 0.07 | 0.03 | 0.05 | 0.04 | 0.09 | 0.04 | 0.07 | 0.14 | 0.06 | 0.02 | 0.11 | 0.07 | 0.02 | 0.02 | 0.06 | 0.06 | 0.02 | 39 | | | | | | | | | | |
| 0.15 | 0.02 | 0.16 | 0.26 | 0.05 | 0.17 | 0.21 | 0.09 | 0.01 | 0.01 | 0.01 | 0.13 | 0.00 | 0.03 | 0.15 | 0.24 | 0.23 | 0.32 | 0.05 | 0.24 | 0.11 | 0.31 | 0.37 | 0.42 | 0.44 | 0.02 | 0.51 | 0.19 | 0.45 | 0.30 | 0.43 | 0.01 | 0.52 | 0.10 | 0.10 | 0.60 | 0.45 | 0.18 | 0.03 | 40 | | | | | | | | | |
| 0.25 | 0.12 | 0.09 | 0.10 | 0.10 | 0.11 | 0.12 | 0.04 | 0.02 | 0.03 | 0.04 | 0.03 | 0.05 | 0.12 | 0.19 | 0.30 | 0.16 | 0.16 | 0.00 | 0.10 | 0.06 | 0.06 | 0.30 | 0.02 | 0.08 | 0.32 | 0.18 | 0.20 | 0.41 | 0.16 | 0.28 | 0.05 | 0.49 | 0.06 | 0.07 | 0.22 | 0.30 | 0.44 | 0.07 | 0.68 | 41 | | | | | | | | |
| 0.09 | 0.02 | 0.15 | 0.26 | 0.15 | 0.25 | 0.11 | 0.01 | 0.11 | 0.06 | 0.16 | 0.02 | 0.08 | 0.06 | 0.29 | 0.33 | 0.24 | 0.31 | 0.04 | 0.24 | 0.05 | 0.19 | 0.23 | 0.24 | 0.26 | 0.01 | 0.47 | 0.57 | 0.45 | 0.51 | 0.65 | 0.11 | 0.67 | 0.07 | 0.07 | 0.51 | 0.70 | 0.42 | 0.04 | 0.67 | 0.76 | 42 | | | | | | | |
| 0.33 | 0.29 | 0.11 | 0.02 | 0.05 | 0.04 | 0.12 | 0.09 | 0.15 | 0.09 | 0.02 | 0.12 | 0.04 | 0.24 | 0.02 | 0.00 | 0.04 | 0.03 | 0.11 | 0.05 | 0.12 | 0.17 | 0.15 | 0.32 | 0.29 | 0.47 | 0.21 | 0.01 | 0.04 | 0.15 | 0.06 | 0.06 | 0.19 | 0.15 | 0.04 | 0.25 | 0.09 | 0.42 | 0.01 | 0.30 | 0.70 | 0.33 | 43 | | | | | | |
| 0.27 | 0.35 | 0.06 | 0.22 | 0.11 | 0.23 | 0.07 | 0.06 | 0.21 | 0.20 | 0.05 | 0.19 | 0.07 | 0.11 | 0.15 | 0.08 | 0.13 | 0.32 | 0.03 | 0.16 | 0.23 | 0.28 | 0.04 | 0.55 | 0.51 | 0.61 | 0.57 | 0.18 | 0.15 | 0.40 | 0.40 | 0.03 | 0.16 | 0.20 | 0.11 | 0.65 | 0.44 | 0.17 | 0.11 | 0.23 | 0.36 | 0.13 | 0.67 | 44 | | | | | |
| 0.26 | 0.35 | 0.07 | 0.23 | 0.13 | 0.21 | 0.06 | 0.04 | 0.19 | 0.17 | 0.08 | 0.21 | 0.08 | 0.13 | 0.13 | 0.06 | 0.13 | 0.32 | 0.06 | 0.16 | 0.25 | 0.30 | 0.01 | 0.55 | 0.51 | 0.63 | 0.58 | 0.19 | 0.12 | 0.40 | 0.40 | 0.02 | 0.17 | 0.25 | 0.18 | 0.66 | 0.45 | 0.19 | 0.11 | 0.23 | 0.37 | 0.12 | 0.67 | 0.98 | 45 | | | | |
| 0.22 | 0.17 | 0.03 | 0.06 | 0.14 | 0.05 | 0.01 | 0.01 | 0.02 | 0.02 | 0.12 | 0.01 | 0.01 | 0.02 | 0.07 | 0.03 | 0.14 | 0.04 | 0.03 | 0.05 | 0.15 | 0.02 | 0.23 | 0.13 | 0.01 | 0.32 | 0.04 | 0.15 | 0.25 | 0.12 | 0.08 | 0.05 | 0.16 | 0.18 | 0.09 | 0.05 | 0.10 | 0.20 | 0.11 | 0.35 | 0.53 | 0.19 | 0.51 | 0.42 | 0.43 | 46 | | | |
| 0.05 | 0.01 | 0.15 | 0.20 | 0.04 | 0.04 | 0.03 | 0.08 | 0.04 | 0.08 | 0.26 | 0.06 | 0.00 | 0.04 | 0.00 | 0.08 | 0.17 | 0.14 | 0.04 | 0.10 | 0.15 | 0.02 | 0.10 | 0.17 | 0.00 | 0.09 | 0.10 | 0.24 | 0.15 | 0.16 | 0.22 | 0.16 | 0.20 | 0.11 | 0.14 | 0.18 | 0.29 | 0.05 | 0.05 | 0.10 | 0.02 | 0.23 | 0.07 | 0.17 | 0.17 | 0.51 | 47 | | |
| 0.04 | 0.10 | 0.18 | 0.28 | 0.03 | 0.21 | 0.03 | 0.00 | 0.09 | 0.05 | 0.24 | 0.14 | 0.06 | 0.02 | 0.15 | 0.15 | 0.27 | 0.34 | 0.07 | 0.24 | 0.19 | 0.24 | 0.06 | 0.47 | 0.37 | 0.30 | 0.48 | 0.34 | 0.19 | 0.37 | 0.49 | 0.08 | 0.40 | 0.17 | 0.08 | 0.56 | 0.51 | 0.12 | 0.04 | 0.42 | 0.16 | 0.49 | 0.17 | 0.51 | 0.49 | 0.32 | 0.79 | 48 | |
| 0.01 | 0.16 | 0.04 | 0.13 | 0.16 | 0.27 | 0.03 | 0.05 | 0.12 | 0.16 | 0.09 | 0.16 | 0.04 | 0.01 | 0.13 | 0.07 | 0.14 | 0.26 | 0.04 | 0.13 | 0.08 | 0.32 | 0.18 | 0.47 | 0.52 | 0.33 | 0.49 | 0.38 | 0.31 | 0.42 | 0.53 | 0.11 | 0.31 | 0.17 | 0.12 | 0.58 | 0.42 | 0.01 | 0.03 | 0.37 | 0.11 | 0.43 | 0.20 | 0.56 | 0.56 | 0.15 | 0.18 | 0.55 | 49 |

The values show Spearman correlation coefficients (absolute values) between repeat lengths (white = 0, red = 1).

**Table S5:** Association of the VNTR lengths with gene expression in CF nasal epithelia (N = 46)

| | *AHRR* | | | *EXOC3* | | | *SLC9A3* | | | *CEP72* | | | *TPPP* | | |
|---|---|---|---|---|---|---|---|---|---|---|---|---|---|---|---|
| **VNTR** | **Beta** | **SE** | **P** | **Beta** | **SE** | **P** | **Beta** | **SE** | **P** | **Beta** | **SE** | **P** | **Beta** | **SE** | **P** |
| 1 | 0.003 | 0.002 | 1.10E-1 | -0.001 | 0.002 | 6.47E-01 | -0.002 | 0.002 | 2.61E-01 | 0.002 | 0.002 | 2.57E-01 | 0.001 | 0.001 | 5.30E-01 |
| 2 | 0.002 | 0.001 | 2.04E-2 | 0.000 | 0.001 | 8.04E-01 | 0.000 | 0.001 | 8.85E-01 | 0.001 | 0.001 | 1.96E-01 | 0.001 | 0.001 | 4.02E-01 |
| 3 | 0.000 | 0.010 | 9.63E-1 | -0.008 | 0.010 | 4.30E-01 | -0.017 | 0.010 | 1.04E-01 | -0.009 | 0.008 | 2.93E-01 | 0.006 | 0.006 | 3.24E-01 |
| 4 | 0.000 | 0.000 | 1.57E-1 | 0.000 | 0.000 | 3.18E-02 | 0.000 | 0.000 | 4.72E-01 | 0.000 | 0.000 | 8.61E-01 | 0.000 | 0.000 | 3.45E-01 |
| 5 | -0.002 | 0.005 | 6.74E-1 | -0.003 | 0.005 | 5.39E-01 | -0.002 | 0.005 | 6.61E-01 | -0.002 | 0.004 | 5.49E-01 | 0.004 | 0.003 | 1.43E-01 |
| 6 | 0.003 | 0.002 | 1.11E-1 | 0.003 | 0.002 | 1.36E-01 | 0.000 | 0.002 | 9.28E-01 | 0.002 | 0.001 | 2.11E-01 | 0.000 | 0.001 | 6.87E-01 |
| 7 | -0.001 | 0.001 | 2.55E-1 | -0.001 | 0.001 | 2.46E-01 | 0.000 | 0.001 | 9.57E-01 | -0.003 | 0.001 | 3.93E-03 | 0.000 | 0.001 | 4.49E-01 |
| 8 | 0.003 | 0.012 | 7.70E-1 | 0.010 | 0.011 | 4.04E-01 | 0.019 | 0.011 | 1.00E-01 | 0.017 | 0.009 | 5.56E-02 | 0.001 | 0.006 | 8.69E-01 |
| 9 | -0.008 | 0.007 | 3.10E-1 | -0.008 | 0.007 | 2.91E-01 | -0.009 | 0.008 | 2.50E-01 | -0.010 | 0.006 | 7.39E-02 | -0.001 | 0.004 | 8.46E-01 |
| 10 | -0.001 | 0.001 | 2.34E-1 | 0.001 | 0.001 | 4.78E-01 | 0.002 | 0.001 | 4.49E-02 | 0.000 | 0.001 | 9.85E-01 | 0.000 | 0.000 | 3.44E-01 |
| 11 | 0.000 | 0.001 | 9.02E-1 | 0.003 | 0.001 | 9.26E-03 | 0.004 | 0.001 | 5.59E-03 | 0.001 | 0.001 | 3.54E-01 | -0.001 | 0.001 | 3.75E-01 |
| 12 | 0.002 | 0.002 | 4.15E-1 | 0.002 | 0.002 | 3.38E-01 | 0.000 | 0.002 | 9.78E-01 | 0.001 | 0.002 | 4.74E-01 | -0.001 | 0.001 | 3.22E-01 |
| 13 | 0.006 | 0.005 | 2.04E-1 | 0.002 | 0.005 | 6.34E-01 | -0.012 | 0.004 | 1.28E-02 | -0.005 | 0.004 | 1.87E-01 | -0.001 | 0.003 | 8.19E-01 |
| 14 | 0.000 | 0.001 | 5.91E-1 | 0.000 | 0.001 | 5.24E-01 | 0.000 | 0.001 | 8.70E-01 | -0.001 | 0.001 | 5.68E-02 | -0.001 | 0.000 | 1.59E-01 |
| 15 | 0.001 | 0.004 | 8.34E-1 | 0.002 | 0.004 | 6.62E-01 | 0.000 | 0.004 | 9.14E-01 | -0.004 | 0.003 | 2.12E-01 | 0.001 | 0.002 | 7.47E-01 |
| 16 | -0.001 | 0.001 | 4.82E-1 | 0.001 | 0.001 | 4.36E-01 | -0.001 | 0.001 | 5.57E-01 | 0.000 | 0.001 | 4.99E-01 | 0.000 | 0.001 | 8.42E-01 |
| 17 | 0.004 | 0.003 | 2.67E-1 | -0.003 | 0.003 | 3.90E-01 | -0.005 | 0.003 | 1.36E-01 | 0.001 | 0.003 | 7.05E-01 | 0.003 | 0.002 | 1.44E-01 |
| 18 | -0.016 | 0.010 | 1.10E-1 | 0.002 | 0.010 | 8.66E-01 | 0.011 | 0.010 | 2.94E-01 | -0.008 | 0.008 | 3.28E-01 | -0.002 | 0.006 | 7.51E-01 |
| 19 | -0.002 | 0.003 | 5.73E-1 | -0.001 | 0.003 | 6.46E-01 | -0.002 | 0.003 | 5.12E-01 | 0.000 | 0.002 | 9.11E-01 | -0.001 | 0.002 | 7.49E-01 |
| 20 | -0.001 | 0.001 | 3.35E-1 | 0.002 | 0.001 | 8.91E-02 | 0.001 | 0.001 | 6.09E-01 | 0.001 | 0.001 | 4.04E-01 | 0.000 | 0.001 | 6.83E-01 |
| 21 | 0.003 | 0.010 | 7.49E-1 | -0.016 | 0.010 | 1.02E-01 | 0.002 | 0.010 | 8.74E-01 | 0.002 | 0.008 | 8.47E-01 | -0.001 | 0.006 | 8.43E-01 |
| 22 | 0.001 | 0.008 | 8.96E-1 | 0.010 | 0.008 | 2.34E-01 | 0.008 | 0.009 | 3.43E-01 | 0.006 | 0.007 | 4.07E-01 | 0.000 | 0.005 | 9.42E-01 |
| 23 | -0.005 | 0.016 | 7.63E-1 | 0.009 | 0.016 | 5.72E-01 | 0.007 | 0.017 | 6.89E-01 | -0.013 | 0.013 | 3.19E-01 | -0.012 | 0.009 | 1.67E-01 |
| 24 | 0.013 | 0.006 | 5.27E-2 | 0.004 | 0.007 | 5.77E-01 | -0.010 | 0.007 | 1.57E-01 | -0.002 | 0.005 | 7.11E-01 | 0.008 | 0.004 | 3.63E-02 |
| 25 | 0.002 | 0.003 | 5.11E-1 | -0.002 | 0.003 | 5.62E-01 | -0.006 | 0.003 | 5.45E-02 | 0.000 | 0.003 | 9.20E-01 | 0.004 | 0.002 | 2.94E-02 |
| 26 | 0.002 | 0.001 | 6.59E-2 | -0.001 | 0.001 | 5.88E-01 | -0.003 | 0.001 | 3.92E-02 | -0.001 | 0.001 | 2.04E-01 | 0.000 | 0.001 | 8.27E-01 |
| 27 | 0.010 | 0.006 | 8.91E-2 | -0.004 | 0.006 | 5.00E-01 | -0.008 | 0.006 | 2.08E-01 | 0.005 | 0.005 | 3.10E-01 | 0.005 | 0.003 | 1.01E-01 |
| 28 | 0.003 | 0.002 | 5.14E-2 | -0.002 | 0.002 | 1.27E-01 | -0.002 | 0.002 | 1.92E-01 | 0.001 | 0.001 | 7.01E-01 | 0.000 | 0.001 | 6.95E-01 |

| | *AHRR* | | | *EXOC3* | | | *SLC9A3* | | | *CEP72* | | | *TPPP* | | |
|---|---|---|---|---|---|---|---|---|---|---|---|---|---|---|---|
| **VNTR** | **Beta** | **SE** | **P** | **Beta** | **SE** | **P** | **Beta** | **SE** | **P** | **Beta** | **SE** | **P** | **Beta** | **SE** | **P** |
| 29 | 0.000 | 0.000 | 5.95E-1 | 0.000 | 0.000 | 7.18E-01 | 0.000 | 0.000 | 7.96E-01 | 0.001 | 0.000 | 1.40E-01 | 0.000 | 0.000 | 6.32E-01 |
| 30 | -0.001 | 0.001 | 3.08E-1 | 0.000 | 0.001 | 7.99E-01 | 0.000 | 0.001 | 6.65E-01 | 0.000 | 0.001 | 5.13E-01 | 0.001 | 0.000 | 2.38E-01 |
| 31 | 0.000 | 0.002 | 8.72E-1 | 0.000 | 0.002 | 8.80E-01 | -0.002 | 0.002 | 2.38E-01 | 0.000 | 0.001 | 8.75E-01 | 0.002 | 0.001 | 6.38E-02 |
| 32 | 0.011 | 0.023 | 6.28E-1 | -0.030 | 0.022 | 1.83E-01 | 0.005 | 0.024 | 8.44E-01 | 0.009 | 0.018 | 6.22E-01 | -0.010 | 0.013 | 4.45E-01 |
| 33 | 0.000 | 0.000 | 1.34E-1 | 0.000 | 0.000 | 7.57E-01 | 0.000 | 0.000 | 5.48E-01 | 0.000 | 0.000 | 3.71E-01 | 0.000 | 0.000 | 8.22E-02 |
| 34 | -0.010 | 0.012 | 3.95E-1 | -0.003 | 0.012 | 8.31E-01 | -0.002 | 0.012 | 8.76E-01 | 0.014 | 0.009 | 1.29E-01 | 0.003 | 0.007 | 6.02E-01 |
| 35 | -0.010 | 0.036 | 7.81E-1 | 0.050 | 0.034 | 1.60E-01 | 0.023 | 0.037 | 5.39E-01 | 0.019 | 0.029 | 5.00E-01 | -0.019 | 0.020 | 3.51E-01 |
| 36 | -0.002 | 0.002 | 2.92E-1 | 0.001 | 0.002 | 5.63E-01 | 0.005 | 0.002 | 4.89E-02 | 0.000 | 0.002 | 9.39E-01 | -0.004 | 0.001 | 1.61E-03 |
| 37 | 0.001 | 0.020 | 9.58E-1 | -0.016 | 0.019 | 4.01E-01 | 0.025 | 0.020 | 2.22E-01 | 0.009 | 0.016 | 5.66E-01 | -0.019 | 0.010 | 8.57E-02 |
| 38 | 0.006 | 0.005 | 2.47E-1 | 0.000 | 0.005 | 9.50E-01 | -0.005 | 0.005 | 3.50E-01 | -0.004 | 0.004 | 3.58E-01 | 0.001 | 0.003 | 5.88E-01 |
| 39 | -0.253 | 1.779 | 8.88E-1 | -2.818 | 1.677 | 1.03E-01 | -0.509 | 1.842 | 7.84E-01 | -0.251 | 1.414 | 8.61E-01 | 0.178 | 0.983 | 8.57E-01 |
| 40 | 0.000 | 0.000 | 6.75E-1 | 0.000 | 0.000 | 3.52E-01 | 0.000 | 0.000 | 5.96E-01 | 0.000 | 0.000 | 2.00E-01 | 0.000 | 0.000 | 1.47E-02 |
| 41 | -0.004 | 0.002 | 1.39E-1 | 0.001 | 0.002 | 8.33E-01 | 0.001 | 0.003 | 6.55E-01 | 0.002 | 0.002 | 2.65E-01 | 0.001 | 0.001 | 3.83E-01 |
| 42 | 0.002 | 0.002 | 2.91E-1 | -0.001 | 0.002 | 6.81E-01 | 0.000 | 0.002 | 9.37E-01 | -0.002 | 0.002 | 2.59E-01 | -0.002 | 0.001 | 9.88E-02 |
| 43 | 0.000 | 0.000 | 8.11E-2 | 0.000 | 0.000 | 8.08E-01 | 0.000 | 0.000 | 1.26E-01 | 0.000 | 0.000 | 3.35E-01 | 0.000 | 0.000 | 5.44E-01 |
| 44 | -0.009 | 0.003 | 1.07E-2 | -0.002 | 0.004 | 6.56E-01 | 0.009 | 0.004 | 1.48E-02 | 0.000 | 0.003 | 9.59E-01 | -0.007 | 0.002 | 3.31E-04 |
| 45 | -0.002 | 0.001 | 3.48E-2 | -0.001 | 0.001 | 5.65E-01 | 0.003 | 0.001 | 2.83E-02 | 0.000 | 0.001 | 8.67E-01 | -0.002 | 0.001 | 2.43E-04 |
| 46 | 0.001 | 0.001 | 4.05E-1 | 0.000 | 0.001 | 7.56E-01 | -0.001 | 0.001 | 5.20E-01 | -0.001 | 0.001 | 1.37E-01 | 0.001 | 0.001 | 2.56E-01 |
| 47 | -0.001 | 0.019 | 9.56E-1 | -0.010 | 0.018 | 5.88E-01 | -0.002 | 0.020 | 9.16E-01 | 0.002 | 0.015 | 8.88E-01 | -0.018 | 0.010 | 8.37E-02 |
| 48 | -0.021 | 0.012 | 8.76E-2 | -0.001 | 0.012 | 9.24E-01 | 0.026 | 0.012 | 4.10E-02 | -0.004 | 0.010 | 6.77E-01 | -0.025 | 0.005 | 4.78E-05 |
| 49 | 0.029 | 0.020 | 1.62E-1 | -0.013 | 0.020 | 5.27E-01 | -0.046 | 0.020 | 2.69E-02 | -0.017 | 0.016 | 2.87E-01 | 0.027 | 0.010 | 1.25E-02 |

Association of the mean length (of two haplotypes) of each VNTR with gene expression was tested using linear regression with sex, RIN, PC1-3, first 7 peer factors and *CD45* expression as covariates in the model.

**Table S6:** Association of the N of CpGs in each VNRT with gene expression in CF nasal epithelia (N = 46)

| | *AHRR* | | | *EXOC3* | | | *SLC9A3* | | | *CEP72* | | | *TPPP* | | |
|---|---|---|---|---|---|---|---|---|---|---|---|---|---|---|---|
| **VNTR** | **Beta** | **SE** | **P** | **Beta** | **SE** | **P** | **Beta** | **SE** | **P** | **Beta** | **SE** | **P** | **Beta** | **SE** | **P** |
| 1 | - | - | - | - | - | - | - | - | - | - | - | - | - | - | - |
| 2 | 0.067 | 0.031 | 4.04E-02 | 0.016 | 0.033 | 6.20E-01 | 0.000 | 0.035 | 9.91E-01 | 0.034 | 0.026 | 2.07E-01 | 0.011 | 0.018 | 5.64E-01 |
| 3 | 0.007 | 0.197 | 9.71E-01 | -0.147 | 0.193 | 4.52E-01 | -0.327 | 0.196 | 1.05E-01 | -0.163 | 0.154 | 3.00E-01 | 0.094 | 0.108 | 3.90E-01 |
| 4 | -0.004 | 0.003 | 1.78E-01 | -0.006 | 0.003 | 4.14E-02 | 0.003 | 0.003 | 4.20E-01 | 0.001 | 0.003 | 8.05E-01 | -0.002 | 0.002 | 3.81E-01 |
| 5 | -0.047 | 0.130 | 7.20E-01 | -0.073 | 0.128 | 5.73E-01 | -0.043 | 0.135 | 7.52E-01 | -0.065 | 0.103 | 5.31E-01 | 0.099 | 0.070 | 1.67E-01 |
| 6 | 0.027 | 0.016 | 1.10E-01 | 0.025 | 0.016 | 1.36E-01 | 0.002 | 0.018 | 9.26E-01 | 0.017 | 0.013 | 2.11E-01 | 0.004 | 0.009 | 6.88E-01 |
| 7 | -0.193 | 0.145 | 1.94E-01 | -0.090 | 0.146 | 5.45E-01 | 0.135 | 0.153 | 3.83E-01 | -0.205 | 0.113 | 7.86E-02 | -0.079 | 0.081 | 3.40E-01 |
| 8 | 0.243 | 0.185 | 1.99E-01 | 0.150 | 0.185 | 4.23E-01 | -0.090 | 0.197 | 6.51E-01 | 0.337 | 0.139 | 2.11E-02 | 0.086 | 0.104 | 4.14E-01 |
| 9 | -0.139 | 0.118 | 2.49E-01 | -0.040 | 0.119 | 7.38E-01 | -0.038 | 0.125 | 7.64E-01 | -0.086 | 0.095 | 3.70E-01 | -0.014 | 0.067 | 8.30E-01 |
| 10 | -0.036 | 0.032 | 2.81E-01 | 0.035 | 0.032 | 2.86E-01 | 0.076 | 0.031 | 2.19E-02 | 0.006 | 0.026 | 8.11E-01 | -0.017 | 0.018 | 3.52E-01 |
| 11 | -0.001 | 0.017 | 9.42E-01 | 0.042 | 0.015 | 1.05E-02 | 0.047 | 0.016 | 5.50E-03 | 0.013 | 0.014 | 3.41E-01 | -0.009 | 0.009 | 3.49E-01 |
| 12 | 0.031 | 0.040 | 4.46E-01 | 0.035 | 0.039 | 3.83E-01 | -0.001 | 0.042 | 9.78E-01 | 0.022 | 0.032 | 4.88E-01 | -0.022 | 0.022 | 3.32E-01 |
| 13 | 0.095 | 0.074 | 2.06E-01 | 0.035 | 0.074 | 6.37E-01 | -0.187 | 0.071 | 1.30E-02 | -0.068 | 0.059 | 2.61E-01 | -0.006 | 0.042 | 8.79E-01 |
| 14 | -0.006 | 0.012 | 6.32E-01 | 0.009 | 0.012 | 4.56E-01 | -0.004 | 0.013 | 7.42E-01 | -0.019 | 0.009 | 5.44E-02 | -0.010 | 0.007 | 1.44E-01 |
| 15 | -0.012 | 0.091 | 8.96E-01 | -0.007 | 0.090 | 9.39E-01 | 0.037 | 0.095 | 6.97E-01 | -0.079 | 0.071 | 2.75E-01 | 0.026 | 0.050 | 6.08E-01 |
| 16 | -0.004 | 0.006 | 5.36E-01 | 0.005 | 0.006 | 4.36E-01 | -0.004 | 0.007 | 5.75E-01 | 0.004 | 0.005 | 4.84E-01 | 0.001 | 0.004 | 7.61E-01 |
| 17 | 0.042 | 0.043 | 3.41E-01 | -0.040 | 0.043 | 3.58E-01 | -0.068 | 0.044 | 1.32E-01 | 0.012 | 0.035 | 7.42E-01 | 0.029 | 0.024 | 2.25E-01 |
| 18 | -0.104 | 0.075 | 1.77E-01 | -0.012 | 0.076 | 8.79E-01 | 0.115 | 0.077 | 1.47E-01 | -0.065 | 0.060 | 2.92E-01 | -0.030 | 0.042 | 4.91E-01 |
| 19 | -0.085 | 0.202 | 6.76E-01 | -0.039 | 0.200 | 8.46E-01 | -0.159 | 0.208 | 4.51E-01 | -0.007 | 0.161 | 9.65E-01 | -0.035 | 0.112 | 7.56E-01 |
| 20 | -0.012 | 0.011 | 2.98E-01 | 0.019 | 0.011 | 8.48E-02 | 0.009 | 0.012 | 4.53E-01 | 0.007 | 0.009 | 4.25E-01 | -0.003 | 0.006 | 6.52E-01 |
| 21 | 0.166 | 0.414 | 6.92E-01 | -0.733 | 0.387 | 6.76E-02 | -0.038 | 0.430 | 9.30E-01 | -0.001 | 0.330 | 9.98E-01 | 0.028 | 0.229 | 9.02E-01 |
| 22 | 0.028 | 0.487 | 9.54E-01 | 0.421 | 0.473 | 3.80E-01 | 0.262 | 0.502 | 6.06E-01 | -0.073 | 0.387 | 8.51E-01 | -0.156 | 0.267 | 5.63E-01 |
| 23 | 0.001 | 0.097 | 9.95E-01 | -0.004 | 0.096 | 9.70E-01 | -0.020 | 0.101 | 8.41E-01 | -0.100 | 0.075 | 1.95E-01 | -0.044 | 0.053 | 4.10E-01 |
| 24 | 0.280 | 0.245 | 2.62E-01 | -0.068 | 0.246 | 7.86E-01 | -0.483 | 0.244 | 5.70E-02 | -0.025 | 0.199 | 9.02E-01 | 0.317 | 0.126 | 1.71E-02 |
| 25 | 0.056 | 0.096 | 5.63E-01 | -0.047 | 0.094 | 6.19E-01 | -0.174 | 0.095 | 7.59E-02 | 0.015 | 0.076 | 8.50E-01 | 0.114 | 0.049 | 2.74E-02 |
| 26 | 0.026 | 0.014 | 6.47E-02 | -0.007 | 0.014 | 6.11E-01 | -0.030 | 0.014 | 3.87E-02 | -0.015 | 0.011 | 2.01E-01 | 0.002 | 0.008 | 8.31E-01 |
| 27 | 0.371 | 0.202 | 7.61E-02 | -0.140 | 0.208 | 5.06E-01 | -0.372 | 0.210 | 8.71E-02 | 0.173 | 0.166 | 3.07E-01 | 0.283 | 0.106 | 1.21E-02 |
| 28 | 0.089 | 0.045 | 5.55E-02 | -0.082 | 0.045 | 7.43E-02 | -0.117 | 0.045 | 1.35E-02 | 0.009 | 0.038 | 8.09E-01 | 0.038 | 0.025 | 1.46E-01 |

| | *AHRR* | | | *EXOC3* | | | *SLC9A3* | | | *CEP72* | | | *TPPP* | | |
|---|---|---|---|---|---|---|---|---|---|---|---|---|---|---|---|
| **VNTR** | **Beta** | **SE** | **P** | **Beta** | **SE** | **P** | **Beta** | **SE** | **P** | **Beta** | **SE** | **P** | **Beta** | **SE** | **P** |
| 29 | -0.030 | 0.085 | 7.30E-01 | -0.087 | 0.082 | 2.96E-01 | 0.024 | 0.088 | 7.85E-01 | -0.023 | 0.067 | 7.36E-01 | -0.013 | 0.047 | 7.88E-01 |
| 30 | -0.016 | 0.014 | 2.59E-01 | -0.003 | 0.014 | 8.25E-01 | -0.004 | 0.014 | 7.66E-01 | -0.007 | 0.011 | 5.02E-01 | 0.008 | 0.007 | 2.73E-01 |
| 31 | -0.027 | 0.044 | 5.39E-01 | 0.027 | 0.044 | 5.41E-01 | -0.038 | 0.046 | 4.11E-01 | -0.005 | 0.035 | 8.92E-01 | 0.036 | 0.024 | 1.38E-01 |
| 32 | -0.017 | 0.243 | 9.45E-01 | -0.116 | 0.238 | 6.30E-01 | -0.335 | 0.245 | 1.80E-01 | 0.014 | 0.193 | 9.44E-01 | 0.225 | 0.128 | 8.95E-02 |
| 33 | -0.003 | 0.002 | 1.23E-01 | -0.001 | 0.002 | 7.91E-01 | -0.001 | 0.002 | 5.98E-01 | 0.001 | 0.002 | 4.37E-01 | 0.002 | 0.001 | 1.04E-01 |
| 34 | -0.226 | 0.183 | 2.27E-01 | 0.087 | 0.184 | 6.39E-01 | 0.064 | 0.194 | 7.45E-01 | 0.192 | 0.145 | 1.95E-01 | -0.025 | 0.104 | 8.11E-01 |
| 35 | - | - | - | - | - | - | - | - | - | - | - | - | - | - | - |
| 36 | -0.022 | 0.020 | 2.98E-01 | 0.012 | 0.020 | 5.70E-01 | 0.041 | 0.020 | 5.22E-02 | -0.002 | 0.016 | 9.10E-01 | -0.033 | 0.010 | 1.68E-03 |
| 37 | 0.036 | 0.458 | 9.38E-01 | -0.364 | 0.446 | 4.21E-01 | 0.555 | 0.464 | 2.41E-01 | 0.211 | 0.362 | 5.65E-01 | -0.431 | 0.241 | 8.37E-02 |
| 38 | 0.242 | 0.281 | 3.97E-01 | -0.394 | 0.271 | 1.57E-01 | -0.430 | 0.285 | 1.41E-01 | -0.060 | 0.226 | 7.93E-01 | 0.276 | 0.149 | 7.40E-02 |
| 39 | - | - | - | - | - | - | - | - | - | - | - | - | - | - | - |
| 40 | 0.000 | 0.006 | 9.96E-01 | 0.005 | 0.005 | 3.73E-01 | -0.002 | 0.006 | 7.14E-01 | 0.006 | 0.004 | 1.62E-01 | 0.007 | 0.003 | 1.90E-02 |
| 41 | -0.120 | 0.078 | 1.32E-01 | 0.017 | 0.079 | 8.33E-01 | 0.039 | 0.083 | 6.42E-01 | 0.070 | 0.063 | 2.73E-01 | 0.040 | 0.044 | 3.73E-01 |
| 42 | 0.034 | 0.042 | 4.18E-01 | -0.018 | 0.042 | 6.67E-01 | 0.007 | 0.044 | 8.77E-01 | -0.038 | 0.033 | 2.55E-01 | -0.045 | 0.022 | 4.74E-02 |
| 43 | 0.007 | 0.004 | 5.97E-02 | 0.001 | 0.004 | 7.84E-01 | -0.006 | 0.004 | 9.99E-02 | -0.003 | 0.003 | 3.67E-01 | 0.002 | 0.002 | 4.62E-01 |
| 44 | -0.971 | 0.471 | 4.77E-02 | -0.263 | 0.492 | 5.97E-01 | 1.097 | 0.482 | 3.00E-02 | -0.106 | 0.399 | 7.93E-01 | -0.949 | 0.219 | 1.44E-04 |
| 45 | -0.030 | 0.013 | 2.52E-02 | -0.008 | 0.014 | 5.43E-01 | 0.031 | 0.013 | 2.63E-02 | -0.001 | 0.011 | 9.05E-01 | -0.025 | 0.006 | 3.26E-04 |
| 46 | 0.008 | 0.010 | 4.41E-01 | -0.003 | 0.010 | 7.78E-01 | -0.006 | 0.011 | 5.64E-01 | -0.012 | 0.008 | 1.31E-01 | 0.006 | 0.006 | 3.09E-01 |
| 47 | - | - | - | - | - | - | - | - | - | - | - | - | - | - | - |
| 48 | -0.401 | 0.438 | 3.68E-01 | -0.409 | 0.431 | 3.50E-01 | 0.296 | 0.457 | 5.22E-01 | -0.209 | 0.351 | 5.57E-01 | -0.653 | 0.216 | 4.93E-03 |
| 49 | 0.133 | 0.092 | 1.59E-01 | -0.104 | 0.092 | 2.66E-01 | -0.206 | 0.092 | 3.19E-02 | -0.063 | 0.075 | 4.08E-01 | 0.125 | 0.048 | 1.31E-02 |

Association of the mean number of CpGs (of two haplotypes) within each VNTR with gene expression was tested using linear regression with sex, RIN, PC1-3, first 7 peer factors and *CD45* expression as covariates in the model.

**Table S7:** Expression levels (TPM) of the genes in the region

| **Gene** | **Min** | **Max** | **Median** |
|---|---|---|---|
| AHRR | 0.0 | 1.1 | 0.3 |
| EXOC3 | 20.8 | 37.3 | 29.8 |
| SLC9A3 | 1.1 | 179.2 | 14.2 |
| CEP72 | 2.8 | 13.7 | 5.6 |
| TPPP | 1.9 | 64.2 | 34.4 |

**Table S8:** Association of the longer and shorter length of the two copies per individual for VNTR #45 and #11 with gene expression compared to average

| VNTR | Gene | Two Copies | β | SE | P | $R^2$ |
|---|---|---|---|---|---|---|
| **#45** | **SLC9A3** | Average | 0.0026 | 0.0011 | 2.83E-02 | 0.08 |
| | | Longer | 0.0010 | 0.0007 | 1.59E-01 | 0.03 |
| | | Shorter | 0.0038 | 0.0014 | 1.36E-02 | 0.09 |
| | **TPPP** | Average | -0.0022 | 0.0005 | 2.43E-04 | 0.05 |
| | | Longer | -0.0011 | 0.0003 | 1.63E-03 | 0.04 |
| | | Shorter | -0.0017 | 0.0008 | 4.00E-02 | 0.01 |
| **#11** | **SLC9A3** | Average | 0.0035 | 0.0012 | 5.59E-03 | 0.12 |
| | | Longer | 0.0032 | 0.0010 | 2.81E-03 | 0.13 |
| | | Shorter | 0.0020 | 0.0011 | 8.94E-02 | 0.04 |

Association of the shorter and longer length (between the two haplotypes) of each VNTR with gene expression was tested using linear regression with sex, RIN, PC1-3, first 7 peer factors and *CD45* expression as covariates in the model.

**Table S9:** Association of VNTRs lengths with *SLC9A3* expression in CF nasal epithelia when all were included in the model

| VNTR | Beta | SE | P |
|---|---|---|---|
| 10 | 0.000 | 0.001 | 0.90 |
| 11 | 0.005 | 0.001 | 6.85E-5 |
| 13 | -0.010 | 0.004 | 0.026 |
| 26 | 0.000 | 0.002 | 0.94 |
| 36 | -0.005 | 0.004 | 0.25 |
| 44 | 0.036 | 0.019 | 0.07 |
| 45 | -0.010 | 0.006 | 0.11 |
| 48 | 0.036 | 0.020 | 0.08 |
| 49 | -0.008 | 0.021 | 0.71 |

Association of the mean length of VNTRs with *SLC9A3* expressions were tested using linear regression. Sex, RIN, PC1-3, first 7 peer factors and *CD45* expression were included as covariates in the model.

**Table S10:** Association of VNTR CpG numbers with *SLC9A3* expression in CF nasal epithelia when all were included in the model

| VNTR | Beta | SE | P |
|---|---|---|---|
| 10 | 0.002 | 0.030 | 0.95 |
| 11 | 0.058 | 0.015 | 9.43E-4 |
| 13 | -0.150 | 0.076 | 0.06 |
| 26 | -0.014 | 0.017 | 0.42 |
| 36 | -0.030 | 0.036 | 0.42 |
| 44 | -2.189 | 3.032 | 0.48 |
| 45 | 0.079 | 0.083 | 0.35 |
| 48 | 0.675 | 0.422 | 0.12 |
| 49 | -0.141 | 0.110 | 0.21 |

Association of the mean VNTR number of CpGs with *SLC9A3* expressions were tested using linear regression. Sex, RIN, PC1-3, first 7 peer factors and *CD45* expression were included as covariates in the model.

**Table S11:** Association of VNTR #45 with gene expressions in GTEx

| | | AHRR | | | EXOC3 | | | SLC9A3 | | | CEP72 | | | TPPP | | |
|---|---|---|---|---|---|---|---|---|---|---|---|---|---|---|---|---|
| **Tissue** | **N** | **Beta** | **SE** | **P** | **Beta** | **SE** | **P** | **Beta** | **SE** | **P** | **Beta** | **SE** | **P** | **Beta** | **SE** | **P** |
| Adipose Subcutaneous | 581 | -0.03 | 0.03 | 3.94E-01 | -0.03 | 0.02 | 3.85E-02 | 0.07 | 0.02 | 2.25E-03 | -0.12 | 0.03 | 1.90E-06 | 0.00 | 0.02 | 9.88E-01 |
| Adipose Visceral Omentum | 469 | 0.05 | 0.03 | 1.77E-01 | -0.04 | 0.02 | 5.66E-02 | 0.11 | 0.03 | 2.07E-04 | -0.06 | 0.02 | 1.48E-02 | 0.04 | 0.02 | 9.76E-02 |
| Adrenal Gland | 233 | -0.01 | 0.05 | 8.26E-01 | -0.04 | 0.04 | 2.70E-01 | 0.13 | 0.05 | 1.44E-02 | -0.16 | 0.04 | 4.14E-04 | 0.00 | 0.03 | 9.76E-01 |
| Artery Aorta | 387 | -0.02 | 0.03 | 6.53E-01 | -0.02 | 0.02 | 4.54E-01 | 0.12 | 0.03 | 4.24E-04 | -0.13 | 0.03 | 4.19E-05 | 0.06 | 0.02 | 1.38E-03 |
| Artery Coronary | 213 | -0.04 | 0.06 | 4.48E-01 | -0.06 | 0.03 | 6.44E-02 | 0.04 | 0.03 | 2.52E-01 | -0.09 | 0.04 | 2.12E-02 | 0.02 | 0.03 | 5.27E-01 |
| Artery Tibial | 584 | -0.07 | 0.03 | 2.65E-02 | -0.04 | 0.01 | 1.34E-02 | 0.14 | 0.03 | 2.75E-07 | -0.08 | 0.02 | 1.24E-04 | 0.05 | 0.02 | 5.06E-03 |
| Brain Amygdala | 129 | 0.01 | 0.04 | 7.55E-01 | -0.08 | 0.05 | 1.54E-01 | 0.05 | 0.05 | 2.80E-01 | -0.01 | 0.04 | 7.52E-01 | 0.04 | 0.03 | 2.18E-01 |
| Brain_Anterior_Cingulate_Cortex_BA24 | 147 | 0.01 | 0.03 | 8.24E-01 | -0.07 | 0.05 | 1.33E-01 | 0.06 | 0.05 | 2.09E-01 | -0.02 | 0.03 | 5.78E-01 | 0.07 | 0.02 | 7.65E-04 |
| Brain Caudate Basal Ganglia | 194 | -0.02 | 0.05 | 7.14E-01 | -0.05 | 0.03 | 1.60E-01 | 0.02 | 0.04 | 6.36E-01 | 0.06 | 0.03 | 1.30E-02 | 0.09 | 0.02 | 6.89E-05 |
| Brain Cerebellar Hemisphere | 175 | -0.05 | 0.05 | 3.75E-01 | -0.08 | 0.04 | 7.40E-02 | 0.06 | 0.04 | 1.17E-01 | -0.04 | 0.03 | 2.51E-01 | 0.14 | 0.03 | 1.57E-04 |
| Brain Cerebellum | 209 | 0.04 | 0.05 | 3.97E-01 | -0.04 | 0.03 | 1.74E-01 | 0.03 | 0.03 | 2.77E-01 | -0.01 | 0.03 | 7.34E-01 | 0.15 | 0.03 | 2.49E-05 |
| Brain Cortex | 205 | -0.01 | 0.05 | 9.10E-01 | -0.01 | 0.03 | 7.53E-01 | 0.04 | 0.05 | 3.85E-01 | 0.07 | 0.03 | 1.01E-02 | 0.04 | 0.02 | 9.78E-03 |
| Brain_Frontal_Cortex_BA9 | 175 | -0.01 | 0.04 | 8.20E-01 | -0.04 | 0.04 | 2.70E-01 | 0.07 | 0.05 | 1.55E-01 | 0.00 | 0.03 | 9.85E-01 | 0.07 | 0.02 | 3.64E-03 |
| Brain Hippocampus | 165 | 0.04 | 0.02 | 1.35E-01 | -0.06 | 0.04 | 1.17E-01 | 0.11 | 0.04 | 1.35E-02 | -0.02 | 0.04 | 6.21E-01 | 0.05 | 0.02 | 9.36E-03 |
| Brain Hypothalamus | 170 | 0.00 | 0.04 | 9.19E-01 | -0.05 | 0.03 | 1.32E-01 | 0.00 | 0.04 | 9.36E-01 | -0.02 | 0.03 | 4.56E-01 | 0.04 | 0.02 | 2.29E-02 |
| Brain Nucleus Accumbens Basal_Ganglia | 202 | 0.01 | 0.04 | 8.86E-01 | 0.00 | 0.03 | 9.30E-01 | 0.08 | 0.04 | 5.75E-02 | 0.00 | 0.03 | 8.72E-01 | 0.11 | 0.02 | 7.12E-07 |
| Brain Putamen Basal Ganglia | 170 | -0.05 | 0.06 | 3.77E-01 | 0.02 | 0.03 | 6.16E-01 | 0.10 | 0.05 | 5.29E-02 | -0.01 | 0.03 | 8.51E-01 | 0.02 | 0.02 | 3.72E-01 |
| Brain_Spinal_cord_cervical_c-1 | 126 | 0.05 | 0.06 | 3.76E-01 | 0.01 | 0.05 | 9.09E-01 | 0.07 | 0.05 | 1.93E-01 | -0.01 | 0.05 | 9.21E-01 | -0.03 | 0.04 | 4.62E-01 |
| Brain Substantia Nigra | 114 | 0.04 | 0.07 | 5.61E-01 | -0.07 | 0.05 | 1.95E-01 | 0.11 | 0.06 | 4.24E-02 | -0.10 | 0.05 | 2.38E-02 | 0.06 | 0.03 | 7.03E-02 |
| Breast Mammary Tissue | 396 | 0.00 | 0.04 | 9.12E-01 | -0.04 | 0.02 | 5.51E-02 | 0.11 | 0.04 | 1.76E-03 | -0.05 | 0.02 | 8.12E-03 | 0.01 | 0.03 | 6.71E-01 |
| Cells Cultured Fibroblasts | 483 | 0.04 | 0.02 | 1.71E-02 | 0.00 | 0.02 | 9.08E-01 | 0.14 | 0.03 | 7.19E-08 | -0.01 | 0.01 | 2.40E-01 | -0.10 | 0.02 | 1.03E-04 |
| Cells EBV-transformed Lymphocytes | 147 | 0.00 | 0.04 | 9.46E-01 | -0.04 | 0.04 | 3.06E-01 | 0.11 | 0.04 | 1.26E-02 | 0.01 | 0.04 | 8.24E-01 | -0.12 | 0.05 | 1.13E-02 |
| Colon Sigmoid | 318 | -0.03 | 0.05 | 5.70E-01 | -0.04 | 0.03 | 1.33E-01 | 0.13 | 0.04 | 2.33E-03 | -0.13 | 0.04 | 7.55E-04 | 0.03 | 0.02 | 2.07E-01 |
| Colon Transverse | 368 | 0.03 | 0.03 | 3.43E-01 | 0.00 | 0.02 | 9.14E-01 | 0.02 | 0.02 | 2.86E-01 | -0.03 | 0.03 | 3.79E-01 | 0.02 | 0.02 | 4.19E-01 |
| Esophagus Gastroesophageal Junction | 330 | 0.08 | 0.04 | 6.62E-02 | -0.05 | 0.02 | 5.11E-02 | 0.13 | 0.04 | 1.00E-03 | -0.05 | 0.03 | 1.25E-01 | 0.02 | 0.02 | 2.88E-01 |
| Esophagus Mucosa | 497 | -0.07 | 0.03 | 9.24E-03 | -0.05 | 0.02 | 5.45E-03 | 0.12 | 0.02 | 8.56E-07 | -0.01 | 0.02 | 4.44E-01 | 0.02 | 0.02 | 2.82E-01 |
| Esophagus Muscularis | 465 | 0.00 | 0.03 | 8.83E-01 | -0.04 | 0.02 | 3.48E-02 | 0.06 | 0.03 | 4.33E-02 | -0.11 | 0.02 | 4.91E-06 | 0.02 | 0.01 | 8.62E-02 |
| Heart Atrial Appendage | 372 | -0.05 | 0.04 | 1.86E-01 | -0.02 | 0.03 | 3.76E-01 | 0.04 | 0.04 | 2.24E-01 | -0.06 | 0.03 | 3.60E-02 | 0.00 | 0.02 | 8.92E-01 |

| | | AHRR | | | EXOC3 | | | SLC9A3 | | | CEP72 | | | TPPP | | |
|---|---|---|---|---|---|---|---|---|---|---|---|---|---|---|---|---|
| **Tissue** | **N** | **Beta** | **SE** | **P** | **Beta** | **SE** | **P** | **Beta** | **SE** | **P** | **Beta** | **SE** | **P** | **Beta** | **SE** | **P** |
| Heart Left Ventricle | 386 | -0.06 | 0.04 | 8.74E-02 | -0.05 | 0.02 | 9.86E-03 | 0.09 | 0.04 | 1.27E-02 | -0.05 | 0.03 | 8.80E-02 | 0.01 | 0.02 | 5.56E-01 |
| Kidney Cortex | 73 | 0.02 | 0.12 | 8.44E-01 | 0.09 | 0.08 | 2.69E-01 | -0.06 | 0.05 | 2.78E-01 | 0.15 | 0.09 | 9.61E-02 | 0.09 | 0.10 | 3.63E-01 |
| Liver | 208 | 0.04 | 0.05 | 4.08E-01 | -0.04 | 0.03 | 1.51E-01 | 0.03 | 0.04 | 5.07E-01 | -0.03 | 0.04 | 3.67E-01 | 0.00 | 0.04 | 9.38E-01 |
| Lung | 515 | -0.05 | 0.02 | 1.24E-02 | -0.06 | 0.02 | 1.50E-03 | 0.08 | 0.03 | 1.09E-03 | -0.07 | 0.02 | 2.38E-03 | -0.03 | 0.02 | 1.39E-01 |
| Minor Salivary Gland | 144 | 0.02 | 0.05 | 7.30E-01 | 0.01 | 0.04 | 7.91E-01 | 0.03 | 0.05 | 5.77E-01 | -0.17 | 0.05 | 2.44E-03 | -0.06 | 0.05 | 2.52E-01 |
| Muscle Skeletal | 706 | -0.01 | 0.03 | 8.13E-01 | -0.01 | 0.01 | 3.66E-01 | 0.11 | 0.03 | 3.18E-05 | -0.02 | 0.02 | 4.43E-01 | 0.01 | 0.02 | 5.85E-01 |
| Nerve Tibial | 532 | 0.01 | 0.03 | 7.65E-01 | -0.02 | 0.02 | 2.37E-01 | 0.03 | 0.02 | 1.38E-01 | -0.11 | 0.02 | 4.24E-06 | 0.04 | 0.02 | 6.19E-02 |
| Ovary | 167 | 0.14 | 0.05 | 1.20E-02 | -0.01 | 0.03 | 6.93E-01 | 0.13 | 0.05 | 2.27E-02 | -0.12 | 0.04 | 1.02E-03 | 0.00 | 0.03 | 9.51E-01 |
| Pancreas | 305 | 0.01 | 0.04 | 7.43E-01 | -0.08 | 0.02 | 4.29E-06 | 0.11 | 0.04 | 7.38E-03 | -0.07 | 0.03 | 3.00E-02 | 0.03 | 0.04 | 4.97E-01 |
| Pituitary | 237 | -0.01 | 0.04 | 8.18E-01 | -0.06 | 0.03 | 3.22E-02 | 0.06 | 0.05 | 1.70E-01 | -0.02 | 0.03 | 5.47E-01 | 0.04 | 0.02 | 7.61E-02 |
| Prostate | 221 | 0.05 | 0.05 | 3.62E-01 | -0.05 | 0.03 | 8.64E-02 | 0.04 | 0.04 | 3.20E-01 | -0.10 | 0.04 | 4.32E-03 | -0.01 | 0.03 | 8.57E-01 |
| Skin Not Sun Exposed Suprapubic | 517 | -0.02 | 0.02 | 4.16E-01 | -0.02 | 0.01 | 1.89E-01 | 0.10 | 0.02 | 1.44E-05 | -0.05 | 0.02 | 3.13E-02 | 0.01 | 0.01 | 6.54E-01 |
| Skin Sun Exposed Lower Leg | 605 | -0.04 | 0.02 | 1.98E-02 | -0.01 | 0.01 | 3.16E-01 | 0.12 | 0.02 | 3.80E-08 | -0.04 | 0.02 | 8.19E-02 | 0.03 | 0.02 | 1.14E-01 |
| Small Intestine Terminal Ileum | 174 | -0.06 | 0.05 | 2.18E-01 | 0.01 | 0.03 | 8.63E-01 | 0.04 | 0.02 | 4.15E-02 | -0.07 | 0.03 | 3.57E-02 | 0.01 | 0.03 | 8.53E-01 |
| Spleen | 227 | 0.01 | 0.04 | 7.70E-01 | -0.05 | 0.03 | 5.69E-02 | 0.13 | 0.05 | 6.84E-03 | -0.03 | 0.04 | 3.82E-01 | -0.05 | 0.04 | 2.28E-01 |
| Stomach | 324 | -0.03 | 0.04 | 3.89E-01 | -0.05 | 0.02 | 2.13E-02 | 0.03 | 0.02 | 1.20E-01 | -0.08 | 0.03 | 4.87E-03 | 0.01 | 0.02 | 6.70E-01 |
| Testis | 322 | -0.05 | 0.03 | 1.02E-01 | -0.01 | 0.03 | 7.38E-01 | 0.06 | 0.04 | 1.29E-01 | -0.13 | 0.03 | 1.55E-06 | -0.09 | 0.04 | 1.98E-02 |
| Thyroid | 574 | -0.02 | 0.02 | 3.63E-01 | -0.04 | 0.01 | 7.19E-03 | 0.07 | 0.02 | 1.11E-03 | -0.05 | 0.02 | 1.58E-03 | -0.03 | 0.02 | 4.10E-02 |
| Uterus | 129 | 0.03 | 0.06 | 6.13E-01 | 0.00 | 0.04 | 9.30E-01 | 0.05 | 0.05 | 3.44E-01 | -0.05 | 0.03 | 1.30E-01 | 0.04 | 0.04 | 2.74E-01 |
| Vagina | 141 | 0.07 | 0.06 | 2.06E-01 | -0.01 | 0.03 | 8.52E-01 | 0.09 | 0.04 | 9.76E-03 | -0.09 | 0.05 | 6.21E-02 | -0.02 | 0.04 | 6.41E-01 |
| Whole Blood | 670 | 0.01 | 0.03 | 7.98E-01 | -0.02 | 0.01 | 1.46E-01 | 0.08 | 0.02 | 1.64E-03 | -0.02 | 0.01 | 2.55E-01 | -0.05 | 0.02 | 2.29E-03 |

Top 5 genotyping principal components; PEER factors determined by sample size (N): 15 factors for $N<150$, 30 factors for $150\leq N<250$, 45 factors for $250\leq N<350$, and 60 factors for $N\geq350$; sequencing platform; sequencing protocol; and sex were included in the model as covariates.

**Table S12:** Association of VNTR #11 with gene expressions in GTEx

| | | AHRR | | | EXOC3 | | | SLC9A3 | | | CEP72 | | | TPPP | | |
|---|---|---|---|---|---|---|---|---|---|---|---|---|---|---|---|---|
| **Tissue** | **N** | **Beta** | **SE** | **P** | **Beta** | **SE** | **P** | **Beta** | **SE** | **P** | **Beta** | **SE** | **P** | **Beta** | **SE** | **P** |
| Adipose Subcutaneous | 581 | -0.01 | 0.03 | 6.41E-01 | 0.01 | 0.02 | 4.34E-01 | -0.20 | 0.02 | 1.70E-19 | 0.05 | 0.03 | 6.25E-02 | 0.02 | 0.02 | 2.04E-01 |
| Adipose Visceral Omentum | 469 | 0.00 | 0.03 | 9.36E-01 | 0.02 | 0.02 | 2.57E-01 | -0.19 | 0.03 | 9.37E-12 | 0.06 | 0.02 | 8.89E-03 | -0.01 | 0.02 | 6.83E-01 |
| Adrenal Gland | 233 | -0.01 | 0.04 | 8.18E-01 | 0.03 | 0.03 | 2.85E-01 | -0.29 | 0.04 | 5.10E-13 | 0.01 | 0.04 | 7.49E-01 | 0.02 | 0.03 | 3.70E-01 |
| Artery Aorta | 387 | -0.01 | 0.04 | 8.73E-01 | 0.04 | 0.02 | 1.25E-01 | -0.21 | 0.03 | 8.38E-10 | 0.05 | 0.03 | 1.04E-01 | 0.01 | 0.02 | 7.45E-01 |
| Artery Coronary | 213 | 0.00 | 0.05 | 9.61E-01 | 0.02 | 0.03 | 4.84E-01 | -0.14 | 0.03 | 2.40E-06 | 0.05 | 0.04 | 2.29E-01 | -0.02 | 0.03 | 5.29E-01 |
| Artery Tibial | 584 | 0.04 | 0.03 | 1.80E-01 | 0.02 | 0.01 | 2.10E-01 | -0.16 | 0.03 | 1.54E-08 | 0.04 | 0.02 | 8.18E-02 | -0.01 | 0.02 | 7.61E-01 |
| Brain Amygdala | 129 | 0.12 | 0.05 | 1.03E-02 | 0.08 | 0.06 | 1.60E-01 | -0.24 | 0.04 | 3.64E-07 | 0.03 | 0.04 | 5.17E-01 | -0.07 | 0.03 | 3.65E-02 |
| Brain Anterior Cingulate Cortex BA24 | 147 | 0.03 | 0.03 | 2.74E-01 | 0.05 | 0.04 | 2.57E-01 | -0.26 | 0.04 | 6.79E-11 | -0.01 | 0.03 | 7.44E-01 | -0.02 | 0.02 | 3.70E-01 |
| Brain Caudate Basal Ganglia | 194 | 0.04 | 0.05 | 3.99E-01 | 0.00 | 0.03 | 9.70E-01 | -0.26 | 0.03 | 5.81E-12 | 0.03 | 0.02 | 1.56E-01 | -0.03 | 0.02 | 1.85E-01 |
| Brain Cerebellar Hemisphere | 175 | 0.05 | 0.05 | 3.04E-01 | 0.04 | 0.04 | 3.40E-01 | -0.11 | 0.03 | 2.02E-03 | 0.02 | 0.03 | 6.29E-01 | -0.02 | 0.03 | 5.97E-01 |
| Brain Cerebellum | 209 | 0.01 | 0.05 | 8.50E-01 | 0.02 | 0.03 | 5.92E-01 | -0.12 | 0.03 | 8.07E-06 | 0.02 | 0.03 | 5.12E-01 | -0.09 | 0.03 | 1.49E-02 |
| Brain Cortex | 205 | -0.02 | 0.04 | 6.17E-01 | 0.04 | 0.03 | 1.66E-01 | -0.29 | 0.03 | 3.03E-13 | 0.02 | 0.02 | 3.30E-01 | -0.03 | 0.01 | 3.54E-02 |
| Brain Frontal Cortex BA9 | 175 | 0.05 | 0.04 | 1.76E-01 | 0.03 | 0.04 | 4.36E-01 | -0.25 | 0.04 | 2.77E-10 | -0.02 | 0.03 | 3.96E-01 | -0.02 | 0.02 | 3.98E-01 |
| Brain Hippocampus | 165 | 0.04 | 0.03 | 1.59E-01 | 0.04 | 0.04 | 2.79E-01 | -0.21 | 0.05 | 2.75E-05 | -0.02 | 0.04 | 6.09E-01 | -0.02 | 0.02 | 3.91E-01 |
| Brain Hypothalamus | 170 | -0.05 | 0.04 | 1.88E-01 | 0.05 | 0.04 | 1.92E-01 | -0.23 | 0.04 | 2.03E-09 | -0.01 | 0.03 | 7.37E-01 | -0.03 | 0.02 | 1.91E-01 |
| Brain Nucleus Accumbens Basal Ganglia | 202 | 0.04 | 0.03 | 2.30E-01 | 0.09 | 0.03 | 3.56E-03 | -0.20 | 0.04 | 2.34E-07 | 0.04 | 0.03 | 8.84E-02 | 0.00 | 0.02 | 9.72E-01 |
| Brain Putamen Basal Ganglia | 170 | -0.02 | 0.05 | 6.57E-01 | -0.02 | 0.03 | 4.52E-01 | -0.25 | 0.04 | 8.49E-10 | 0.02 | 0.03 | 5.17E-01 | -0.05 | 0.02 | 1.18E-02 |
| Brain Spinal Cord Cervical c-1 | 126 | 0.00 | 0.06 | 9.46E-01 | -0.02 | 0.05 | 6.86E-01 | -0.20 | 0.05 | 4.93E-05 | 0.06 | 0.05 | 2.78E-01 | -0.04 | 0.04 | 3.19E-01 |
| Brain Substantia Nigra | 114 | 0.05 | 0.06 | 3.94E-01 | 0.03 | 0.04 | 5.27E-01 | -0.16 | 0.04 | 6.06E-04 | -0.01 | 0.04 | 7.99E-01 | 0.03 | 0.03 | 2.90E-01 |
| Breast Mammary Tissue | 396 | -0.05 | 0.04 | 2.44E-01 | 0.00 | 0.02 | 8.70E-01 | -0.28 | 0.03 | 7.20E-14 | 0.01 | 0.02 | 5.76E-01 | 0.05 | 0.03 | 8.99E-02 |
| Cells Cultured Fibroblasts | 483 | -0.02 | 0.02 | 1.69E-01 | 0.03 | 0.02 | 3.14E-02 | -0.23 | 0.02 | 1.13E-19 | 0.01 | 0.01 | 5.23E-01 | 0.04 | 0.03 | 9.40E-02 |
| Cells EBV-transformed Lymphocytes | 147 | -0.01 | 0.05 | 9.02E-01 | 0.05 | 0.05 | 3.33E-01 | -0.18 | 0.05 | 6.08E-04 | -0.05 | 0.04 | 2.52E-01 | 0.03 | 0.06 | 5.79E-01 |
| Colon Sigmoid | 318 | 0.03 | 0.04 | 4.95E-01 | 0.02 | 0.02 | 4.44E-01 | -0.16 | 0.04 | 1.97E-05 | 0.00 | 0.03 | 9.80E-01 | 0.00 | 0.02 | 8.35E-01 |
| Colon Transverse | 368 | 0.03 | 0.03 | 2.87E-01 | 0.04 | 0.02 | 3.30E-02 | -0.06 | 0.02 | 3.67E-05 | 0.03 | 0.03 | 2.02E-01 | 0.01 | 0.02 | 3.89E-01 |
| Esophagus Gastroesophageal Junction | 330 | -0.01 | 0.04 | 8.69E-01 | 0.04 | 0.02 | 8.82E-02 | -0.16 | 0.04 | 2.16E-05 | 0.01 | 0.03 | 7.46E-01 | -0.02 | 0.02 | 4.20E-01 |
| Esophagus Mucosa | 497 | -0.03 | 0.03 | 2.08E-01 | 0.02 | 0.02 | 3.47E-01 | -0.13 | 0.02 | 3.03E-08 | 0.03 | 0.02 | 1.03E-01 | 0.01 | 0.02 | 6.27E-01 |
| Esophagus Muscularis | 465 | 0.04 | 0.03 | 2.21E-01 | 0.06 | 0.02 | 1.12E-03 | -0.20 | 0.02 | 1.32E-14 | 0.01 | 0.02 | 5.88E-01 | -0.02 | 0.01 | 1.76E-01 |
| Heart Atrial Appendage | 372 | 0.02 | 0.03 | 6.06E-01 | 0.03 | 0.02 | 2.20E-01 | -0.23 | 0.03 | 1.04E-14 | 0.06 | 0.02 | 1.37E-02 | -0.01 | 0.02 | 5.71E-01 |

| | | AHRR | | | EXOC3 | | | SLC9A3 | | | CEP72 | | | TPPP | | |
|---|---|---|---|---|---|---|---|---|---|---|---|---|---|---|---|---|
| **Tissue** | **N** | **Beta** | **SE** | **P** | **Beta** | **SE** | **P** | **Beta** | **SE** | **P** | **Beta** | **SE** | **P** | **Beta** | **SE** | **P** |
| Heart Left Ventricle | 386 | 0.01 | 0.03 | 8.42E-01 | 0.04 | 0.02 | 1.93E-02 | -0.22 | 0.03 | 9.20E-12 | 0.03 | 0.03 | 2.44E-01 | 0.01 | 0.01 | 5.38E-01 |
| Kidney Cortex | 73 | 0.06 | 0.07 | 4.18E-01 | 0.01 | 0.05 | 7.93E-01 | -0.02 | 0.03 | 5.21E-01 | 0.10 | 0.06 | 8.30E-02 | 0.00 | 0.06 | 9.65E-01 |
| Liver | 208 | 0.01 | 0.05 | 8.13E-01 | -0.05 | 0.03 | 1.39E-01 | -0.19 | 0.04 | 3.45E-06 | -0.06 | 0.04 | 1.55E-01 | 0.03 | 0.04 | 5.35E-01 |
| Lung | 515 | 0.01 | 0.02 | 5.84E-01 | 0.03 | 0.02 | 1.27E-01 | -0.18 | 0.02 | 9.80E-13 | 0.03 | 0.02 | 1.68E-01 | -0.03 | 0.02 | 6.71E-02 |
| Minor Salivary Gland | 144 | 0.00 | 0.05 | 9.85E-01 | 0.03 | 0.04 | 4.28E-01 | -0.17 | 0.04 | 9.15E-05 | 0.01 | 0.06 | 7.97E-01 | 0.00 | 0.05 | 9.36E-01 |
| Muscle Skeletal | 706 | -0.01 | 0.03 | 8.10E-01 | 0.04 | 0.01 | 5.09E-03 | -0.20 | 0.02 | 6.87E-17 | 0.02 | 0.02 | 3.40E-01 | -0.02 | 0.02 | 2.75E-01 |
| Nerve Tibial | 532 | -0.01 | 0.03 | 8.46E-01 | -0.02 | 0.02 | 3.43E-01 | -0.01 | 0.02 | 7.12E-01 | 0.02 | 0.02 | 4.08E-01 | -0.01 | 0.02 | 6.31E-01 |
| Ovary | 167 | 0.07 | 0.05 | 1.52E-01 | 0.04 | 0.02 | 8.11E-02 | -0.29 | 0.04 | 9.68E-11 | 0.04 | 0.03 | 1.83E-01 | -0.03 | 0.03 | 2.71E-01 |
| Pancreas | 305 | -0.03 | 0.04 | 4.12E-01 | 0.00 | 0.02 | 9.71E-01 | -0.24 | 0.03 | 4.34E-11 | 0.05 | 0.03 | 1.43E-01 | 0.00 | 0.04 | 9.91E-01 |
| Pituitary | 237 | 0.05 | 0.03 | 1.37E-01 | -0.02 | 0.03 | 4.14E-01 | -0.24 | 0.04 | 2.21E-08 | 0.02 | 0.03 | 4.58E-01 | 0.00 | 0.02 | 8.47E-01 |
| Prostate | 221 | 0.11 | 0.05 | 2.89E-02 | 0.02 | 0.03 | 5.60E-01 | -0.15 | 0.04 | 9.82E-05 | 0.00 | 0.04 | 8.95E-01 | 0.02 | 0.03 | 4.71E-01 |
| Skin Not Sun Exposed Suprapubic | 517 | 0.03 | 0.02 | 2.32E-01 | 0.00 | 0.01 | 9.72E-01 | -0.15 | 0.02 | 4.44E-13 | 0.04 | 0.02 | 7.57E-02 | -0.02 | 0.01 | 1.90E-01 |
| Skin Sun Exposed Lower leg | 605 | 0.00 | 0.02 | 8.53E-01 | 0.02 | 0.01 | 1.83E-01 | -0.15 | 0.02 | 1.10E-13 | 0.03 | 0.02 | 1.51E-01 | 0.03 | 0.02 | 8.75E-02 |
| Small Intestine Terminal Ileum | 174 | -0.06 | 0.05 | 2.41E-01 | 0.03 | 0.03 | 2.99E-01 | 0.00 | 0.02 | 8.45E-01 | -0.03 | 0.03 | 4.09E-01 | 0.00 | 0.04 | 9.26E-01 |
| Spleen | 227 | -0.01 | 0.04 | 8.91E-01 | 0.04 | 0.03 | 1.12E-01 | -0.31 | 0.04 | 7.17E-11 | -0.03 | 0.04 | 3.94E-01 | 0.05 | 0.04 | 2.08E-01 |
| Stomach | 324 | -0.02 | 0.04 | 5.11E-01 | -0.02 | 0.02 | 4.31E-01 | -0.03 | 0.02 | 1.48E-01 | 0.01 | 0.03 | 6.63E-01 | -0.02 | 0.02 | 4.14E-01 |
| Testis | 322 | -0.03 | 0.03 | 3.90E-01 | -0.01 | 0.03 | 7.92E-01 | -0.21 | 0.03 | 2.75E-11 | 0.04 | 0.03 | 1.61E-01 | -0.01 | 0.04 | 7.13E-01 |
| Thyroid | 574 | -0.02 | 0.02 | 4.43E-01 | 0.01 | 0.01 | 3.35E-01 | -0.19 | 0.02 | 2.05E-18 | 0.03 | 0.02 | 1.22E-01 | 0.00 | 0.02 | 9.60E-01 |
| Uterus | 129 | -0.01 | 0.06 | 9.02E-01 | 0.05 | 0.03 | 1.55E-01 | -0.24 | 0.04 | 1.34E-07 | 0.04 | 0.03 | 2.35E-01 | 0.03 | 0.03 | 4.55E-01 |
| Vagina | 141 | -0.01 | 0.06 | 9.26E-01 | 0.04 | 0.03 | 1.87E-01 | -0.15 | 0.03 | 3.10E-05 | 0.03 | 0.05 | 4.92E-01 | -0.06 | 0.04 | 1.28E-01 |
| Whole Blood | 670 | -0.04 | 0.03 | 1.62E-01 | 0.03 | 0.01 | 2.58E-03 | -0.12 | 0.02 | 5.56E-07 | 0.00 | 0.01 | 9.24E-01 | 0.03 | 0.02 | 1.34E-01 |

Top 5 genotyping principal components; PEER factors determined by sample size (N): 15 factors for $N<150$, 30 factors for $150 \leq N<250$, 45 factors for $250 \leq N<350$, and 60 factors for $N \geq 350$; sequencing platform; sequencing protocol; and sex were included in the model as covariates.

**Table S13:** Association of VNTRs overlapping with VNTR #11 and 45 with gene expression in GTEx

| VNTR | Overlapping VNTR coordinate | Gene | GTEx tissue | R-value | p-value | FDR |
|---|---|---|---|---|---|---|
| 11 | chr5:474086-474868 | *SLC9A3* | Adipose Subcutaneous | -0.591 | 5.03E-23 | 2.07E-18 |
| 11 | chr5:474086-474868 | *SLC9A3* | Adipose Visceral Omentum | -0.615 | 1.96E-25 | 1.25E-20 |
| 11 | chr5:474086-474868 | *SLC9A3* | Adrenal Gland | -0.636 | 6.81E-13 | 6.07E-08 |
| 11 | chr5:474086-474868 | *SLC9A3* | Artery Aorta | -0.638 | 9.24E-19 | 6.69E-14 |
| 11 | chr5:474086-474868 | *SLC9A3* | Artery Coronary | -0.429 | 1.19E-05 | 0.091111 |
| 11 | chr5:474086-474868 | *SLC9A3* | Artery Tibial | -0.494 | 1.86E-15 | 3.29E-11 |
| 11 | chr5:474086-474868 | *SLC9A3* | Brain Anterior cingulate cortex BA24 | -0.687 | 2.67E-12 | 3.04E-07 |
| 11 | chr5:474086-474868 | *SLC9A3* | Brain Caudate basal ganglia | -0.618 | 1.92E-11 | 1.24E-06 |
| 11 | chr5:474086-474868 | *SLC9A3* | Brain Cortex | -0.611 | 1.66E-10 | 7.74E-06 |
| 11 | chr5:474086-474868 | *SLC9A3* | Brain Frontal Cortex BA9 | -0.737 | 2.12E-15 | 3.55E-10 |
| 11 | chr5:474086-474868 | *SLC9A3* | Brain Hippocampus | -0.604 | 1.58E-08 | 0.000932 |
| 11 | chr5:474086-474868 | *SLC9A3* | Brain Hypothalamus | -0.594 | 3.13E-08 | 0.001268 |
| 11 | chr5:474086-474868 | *SLC9A3* | Brain Nucleus accumbens basal ganglia | -0.596 | 1.74E-09 | 8.12E-05 |
| 11 | chr5:474086-474868 | *SLC9A3* | Brain Putamen basal ganglia | -0.638 | 3.45E-10 | 2.55E-05 |
| 11 | chr5:474086-474868 | *SLC9A3* | Brain Spinal cord cervical c-1 | -0.698 | 8.16E-10 | 8.51E-05 |
| 11 | chr5:474086-474868 | *SLC9A3* | Breast Mammary Tissue | -0.564 | 1.81E-14 | 7.37E-10 |
| 11 | chr5:474086-474868 | *SLC9A3* | Cells EBV-transformed lymphocytes | -0.588 | 9.58E-07 | 0.032441 |
| 11 | chr5:474086-474868 | *SLC9A3* | Cells Transformed fibroblasts | -0.653 | 2.09E-17 | 9.12E-13 |
| 11 | chr5:474086-474868 | *SLC9A3* | Colon Sigmoid | -0.508 | 3.17E-11 | 7.88E-07 |
| 11 | chr5:474086-474868 | *SLC9A3* | Esophagus Gastroesophageal Junction | -0.453 | 3.71E-09 | 4.40E-05 |
| 11 | chr5:474086-474868 | *SLC9A3* | Esophagus Mucosa | -0.521 | 7.73E-16 | 1.84E-11 |
| 11 | chr5:474086-474868 | *SLC9A3* | Esophagus Muscularis | -0.532 | 1.02E-15 | 2.08E-11 |
| 11 | chr5:474086-474868 | *SLC9A3* | Heart Atrial Appendage | -0.568 | 4.18E-17 | 1.63E-12 |
| 11 | chr5:474086-474868 | *SLC9A3* | Heart Left Ventricle | -0.553 | 7.36E-14 | 3.59E-09 |
| 11 | chr5:474086-474868 | *SLC9A3* | Liver | -0.595 | 2.11E-10 | 1.34E-05 |
| 11 | chr5:474086-474868 | *SLC9A3* | Lung | -0.614 | 1.24E-24 | 7.70E-20 |

| VNTR | Overlapping VNTR coordinate | Gene | GTEx tissue | R-value | p-value | FDR |
|---|---|---|---|---|---|---|
| 11 | chr5:474086-474868 | *SLC9A3* | Minor Salivary Gland | -0.576 | 6.52E-07 | 0.066534 |
| 11 | chr5:474086-474868 | *SLC9A3* | Muscle Skeletal | -0.541 | 7.02E-25 | 3.38E-20 |
| 11 | chr5:474086-474868 | *SLC9A3* | Ovary | -0.782 | 1.30E-16 | 5.47E-11 |
| 11 | chr5:474086-474868 | *SLC9A3* | Pancreas | -0.559 | 4.89E-12 | 1.72E-07 |
| 11 | chr5:474086-474868 | *SLC9A3* | Pituitary | -0.636 | 1.11E-13 | 8.27E-09 |
| 11 | chr5:474086-474868 | *SLC9A3* | Prostate | -0.563 | 1.21E-07 | 0.004585 |
| 11 | chr5:474086-474868 | *SLC9A3* | Skin Not Sun Exposed Suprapubic | -0.555 | 2.13E-21 | 8.21E-17 |
| 11 | chr5:474086-474868 | *SLC9A3* | Skin Sun Exposed Lower leg | -0.471 | 5.97E-16 | 1.07E-11 |
| 11 | chr5:474086-474868 | *SLC9A3* | Spleen | -0.641 | 4.39E-12 | 4.16E-07 |
| 11 | chr5:474086-474868 | *SLC9A3* | Testis | -0.650 | 2.63E-17 | 2.19E-12 |
| 11 | chr5:474086-474868 | *SLC9A3* | Thyroid | -0.624 | 1.21E-28 | 8.02E-24 |
| 11 | chr5:474086-474868 | *SLC9A3* | Uterus | -0.703 | 3.15E-09 | 0.000882 |
| 11 | chr5:474086-474868 | *SLC9A3* | Whole Blood | -0.438 | 3.18E-10 | 3.21E-06 |
| 45 | chr5:662835-663227 | *CEP72* | Adipose Subcutaneous | -0.268 | 3.77E-05 | 0.059043 |
| 45 | chr5:662835-663227 | *CEP72* | Artery Tibial | -0.277 | 2.07E-05 | 0.035203 |
| 45 | chr5:662835-663227 | *CEP72* | Esophagus Muscularis | -0.337 | 1.35E-06 | 0.004492 |
| 45 | chr5:662835-663227 | *CEP72* | Nerve Tibial | -0.276 | 4.23E-05 | 0.053011 |
| 45 | chr5:662835-663227 | *CEP72* | Testis | -0.478 | 5.73E-09 | 6.40E-05 |
| 45 | chr5:662835-663227 | *TPPP* | Brain Cerebellar Hemisphere | 0.511 | 5.82E-07 | 0.007853 |
| 45 | chr5:662835-663227 | *TPPP* | Brain Cerebellum | 0.510 | 4.98E-08 | 0.00058 |
| 45 | chr5:662835-663227 | *TPPP* | Brain Nucleus accumbens basal ganglia | 0.480 | 3.42E-06 | 0.043186 |

**Table S14:** Association of VNTRs overlapping with VNTR #11 and 45 with DNA methylation in PGCG [1]

| VNTR | Overlapping VNTR | CpG probe ID | CpG position | p-value | R-value | Bonferroni adjusted p-value |
|---|---|---|---|---|---|---|
| 11 | chr5:474086-474868 | cg21743597 | 475090 | 3.57E-13 | -0.451 | 3.89E-07 |
| 11 | chr5:474086-474868 | cg21159260 | 475774 | 4.31E-21 | -0.566 | 4.71E-15 |
| 11 | chr5:474086-474868 | cg17233264 | 476290 | 6.30E-11 | -0.410 | 6.87E-05 |
| 45 | chr5:662835-663227 | cg19576778 | 654910 | 1.87E-39 | -0.729 | 2.04E-33 |
| 45 | chr5:662835-663227 | cg16624210 | 671319 | 1.33E-10 | 0.407 | 0.00014 |

**Figure S1:** Association of SNPs with lung function in CF

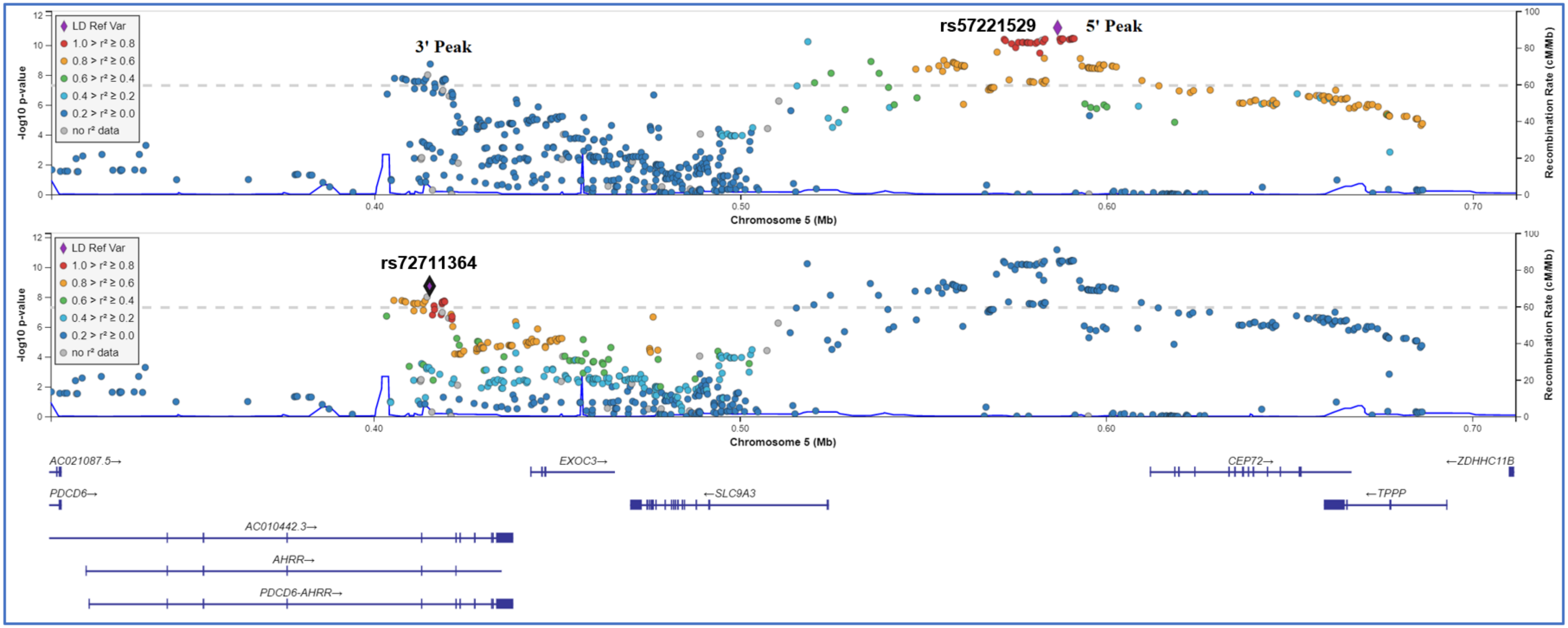


The left y-axis shows SNP p-values for association with lung function in CF [2]. The right y-axis shows estimated recombination rates. The x-axis shows SNP genomic position (HG38). The LD measures are based on EUR population, the index SNP is shown in purple diamond (top associated SNP in 5' peak (top) and 3' peak (bottom)). The plot was created using LocusZoom (https://my.locuszoom.org/).

Rec: Recombination

**Figure S2:** Pair-wise LD between the SNPs associated with lung function in CF patients that were mQTL for the CpGs associated with *SLC9A3* expression in the eQTM analysis in the nasal tissue

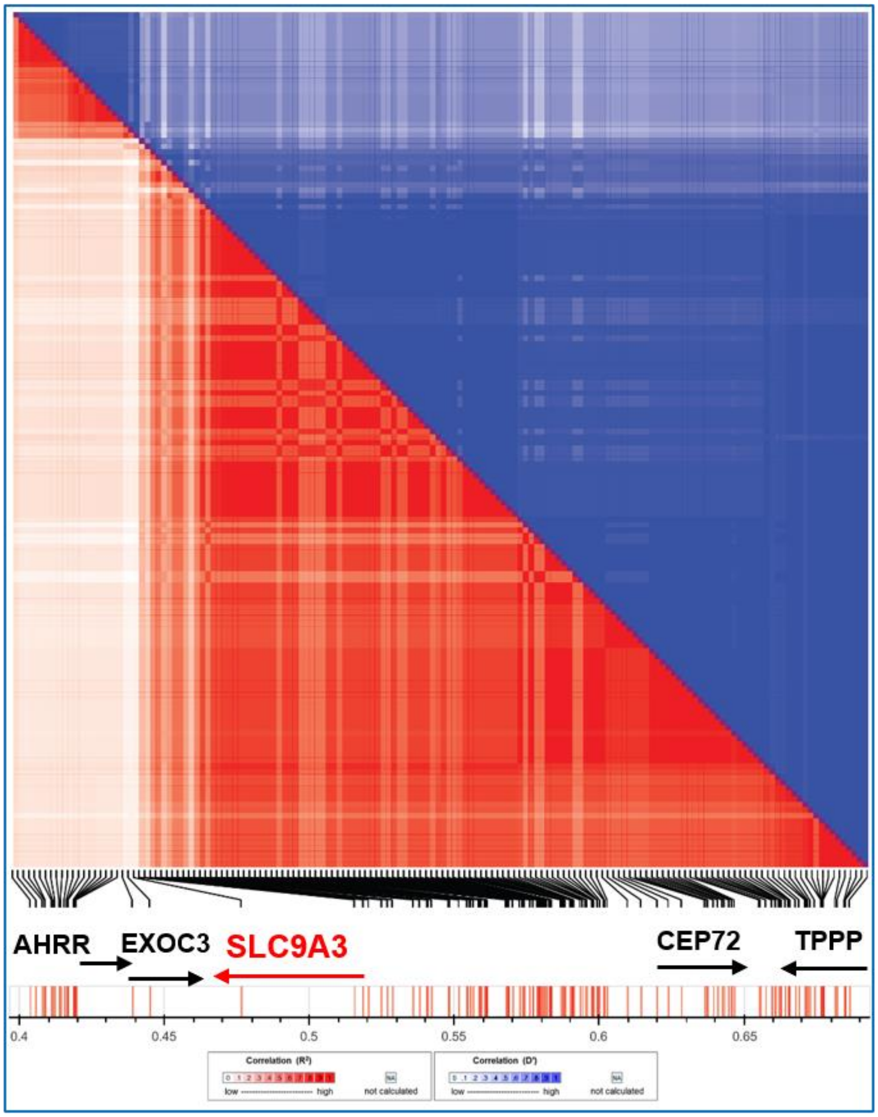


The plot was made using LDMatrix tool from LDLink (https://ldlink.nci.nih.gov/) with European CEU population as reference.
